# Prescribing of glucagon-like peptide-1 receptor agonists for type 2 diabetes by severe mental illness status: a UK primary care cohort study, 2005-2025

**DOI:** 10.64898/2026.11.27.26369133

**Authors:** Stephanie M. Wu, Sophie Picton, Alvin Richards-Belle, Naomi Launders, David P.J. Osborn, Joseph F. Hayes

## Abstract

**Background:** People diagnosed with severe mental illness (SMI) have higher rates of type 2 diabetes mellitus (T2DM), but GLP-1 receptor agonist (GLP-1RA) prescribing in this population remains poorly understood.

**Objective:** To describe GLP-1RA prescribing for T2DM in UK primary care by SMI status.

**Methods:** Using Clinical Practice Research Datalink data (2005–2025), we described GLP-1RA initiators, incidence, prevalence, treatment-pathway position, discontinuation and prescribing continuity. Among adults with incident T2DM, logistic regression compared receipt of a GLP-1RA versus another antidiabetic at each treatment intensification, by SMI status.

**Findings:** Among 1,145,907 adults with incident T2DM, 320,751 (27.9%) had SMI and 102,041 (8.9%) initiated GLP-1RAs. Initiators had high metabolic burden (median HbA1c 71 mmol/mol, BMI 36.5 kg/m²). Although crude prescribing was higher in SMI overall, associations differed by subtype. At third-line intensification, adjusted odds of GLP-1RA initiation were higher in severe depression (OR 1.16, 95% CI 1.13–1.20) but lower in schizophrenia (OR 0.72, 95% CI 0.62– 0.84) and other psychosis (OR 0.82, 95% CI 0.69–0.96), compared to no SMI. Over half discontinued after initiation, around 60% of whom later restarted. Prescribing continuity was highest in bipolar disorder and schizophrenia.

**Conclusions:** Higher overall prescribing in SMI masked lower GLP-1RA initiation among people with schizophrenia or other psychosis despite substantial cardiometabolic risk.

**Clinical implications:** Services should routinely assess GLP-1RA eligibility in people with SMI, particularly those with psychotic disorders, and monitor prescribing across SMI subtypes as eligibility expands. Shared protocols across primary care, diabetes and mental health services may support equitable initiation and sustained treatment.

**Key messages:** *What is already known on this topic:* People with severe mental illness (SMI) have markedly higher rates of type 2 diabetes and cardiovascular disease and face inequities in physical healthcare. GLP-1 receptor agonists (GLP-1RAs) improve cardiometabolic outcomes, including in small trials in schizophrenia, but we do not know whether people with SMI receive them equitably in routine care.

*What this study adds:* Among over one million UK adults with incident type 2 diabetes followed for up to 20 years, crude GLP-1RA prescribing was higher for patients diagnosed with SMI, but this was driven by the large severe depression group and higher metabolic need. After adjustment, people with schizophrenia or other psychosis were less likely to be prescribed a GLP-1RA at treatment intensification and intensified treatment more slowly, despite their higher metabolic burden. Continuity of prescribing after initiation was not lower in SMI and was highest in bipolar disorder and schizophrenia.

*How this study might affect research, practice or policy:* Pooled SMI estimates can obscure inequities affecting people with psychosis. As GLP-1RA eligibility expands, services should examine differences by SMI subtype throughout the prescribing pathway and embed routine GLP-1RA eligibility review within cardiometabolic care for people with psychosis.

## Background

Glucagon-like peptide-1 receptor agonists (GLP-1RAs) are an increasingly important treatment for type 2 diabetes mellitus (T2DM) and obesity. They were licensed in the United Kingdom (UK) for T2DM in 2007 and for obesity in 2022 (NICE, 2022). Beyond lowering blood glucose, they promote weight loss, improve serum lipids, and confer cardiovascular and renal protection (Liu, 2024). These benefits may be particularly relevant to people diagnosed with severe mental illness (SMI; schizophrenia, other psychosis, bipolar disorder and severe depression), who have around 85% higher risk of T2DM than the general population (Vancampfort et al., 2016) and reduced life expectancy, primarily driven by cardiometabolic disease. Trials support the metabolic benefits of GLP-1RAs in antipsychotic-treated patients diagnosed with schizophrenia (Larsen et al., 2017; Ganeshalingam et al., 2025; Siskind et al., 2025), but descriptive studies of routine prescribing are needed to assess equity of care and real-world effectiveness across the SMI spectrum. these small trials do not tell us how GLP-1RAs are prescribed across the broader SMI population in routine care.

People with SMI experience well-documented inequities in the detection and treatment of physical illness, including gaps in access to metformin (Farache Trajano et al., 2025), attributed to diagnostic overshadowing, stigma, socioeconomic disadvantage and fragmentation between mental and physical health services (Rodgers et al., 2018; Bellass et al., 2021). Access to GLP-1RAs may be particularly challenging given their cost, global supply shortages and restrictive eligibility criteria: 2022 NICE guidance recommended GLP-1RAs only after an inadequate response to three oral agents and when treatment of obesity would be beneficial (NICE, 2022; Supplementary Table 1). UK primary care prescribing has largely followed this guidance, restricting GLP-1RAs to a small subset of people with difficult-to-manage T2DM (Ulrich et al., 2026). Whether this holds for people with SMI remains unknown. A small Danish cohort with data until 2015 found higher early GLP-1RA use among people with SMI (Bakkedal et al., 2023), whereas a recent meta-analysis found that people with mental disorders, broadly defined, had poorer diabetes monitoring and were less likely to receive GLP-1RAs (Wagner et al., 2026).

GLP-1RA prescribing is also socially patterned and changing rapidly. In Sweden, prevalence increased fourfold between 2019 and 2023, with a shift towards younger and female recipients (Fors et al., 2025); in the United States (US), newer glucose-lowering drugs are more commonly used by people with higher income and education (Mittman et al., 2024). Initiation may also not translate into sustained treatment: substantial first-year discontinuation, with frequent reinitiation, has been documented in the US (Rodriguez et al., 2025).

## Objective

Using 20 years of UK primary care data, we aimed to characterise GLP-1RA prescribing for T2DM in people with and without SMI. Our objectives were to: (1) describe the sociodemographic and clinical characteristics of GLP-1RA initiators by SMI subtype; (2) examine incidence and point prevalence over time; (3) assess whether SMI status was associated with GLP-1RA initiation at treatment intensification, accounting for sociodemographic and diabetes progression factors; (4) describe GLP-1RA prescribing within the T2DM treatment pathway; and (5) measure discontinuation and continuity of prescribing after initiation.

## Methods

### Study design and data source

We conducted a retrospective cohort study using data recorded from 1 January 2005 to 5 May 2025 in the June 2025 builds of the Clinical Practice Research Datalink (CPRD) Gold and Aurum databases. These databases contain anonymised longitudinal primary care records for over 62 million people and are broadly representative of the UK population (Wolf et al., 2019). For practices located in England, records were linked at the individual level to Hospital Episode Statistics (HES) and Office of National Statistics (ONS) mortality data to supplement ethnicity and death recording, respectively, and at the individual or practice level to 2019 Index of Multiple Deprivation for area-level socioeconomic context. Reporting follows the RECORD-PE guidelines (Langan et al., 2018). Code lists and analysis code are available at https://github.com/smwu/SMI_GLP.

### Participants

The eligible T2DM cohort included adults aged 18–100 with at least 365 days of research-quality registration. T2DM was defined by a medical code or antidiabetic prescription and confirmed by at least one recording of HbA1c ≥48 mmol/mol or fasting plasma glucose *≥*7.0 mmol/L at any time (NICE, 2025; Jeffery et al., 2024). We excluded people initiating insulin within six months of diagnosis (possible type 1 or advanced diabetes) and those whose diabetes records were limited to pregnancy (possible gestational diabetes) (Tate et al., 2017; Jeffery et al., 2024). Follow-up ran from the latest of 1 January 2005, age 18, T2DM diagnosis and 365 days of research-quality registration, until the earliest of 5 May 2025, age 100, deregistration, last data collection or death. The full T2DM cohort included individuals with T2DM recorded before or during study entry, while the incident T2DM cohort comprised only individuals whose first T2DM record occurred after the start of follow-up. Full definitions are provided in the Supplementary Methods, and cohort selection is shown in Supplementary Figure 2.

### Measures

### Severe mental illness

SMI was identified from primary care records as schizophrenia, bipolar disorder, other psychotic disorder, or severe depression. Severe depression required a major depressive disorder code alongside an ever-recorded antidepressant prescription. The diagnosis date was the earlier of the first diagnostic code or the first relevant psychotropic prescription. SMI status used information recorded on or before the relevant date (e.g., T2DM diagnosis or specified timepoint). When more than one diagnostic subtype was recorded, a time-updated hierarchy was applied (schizophrenia, then bipolar disorder, other psychosis, and depression; Supplementary Methods).

### GLP-1RA medication prescription

The outcome was a primary care prescription for any GLP-1RA (albiglutide, dulaglutide, exenatide, liraglutide, lixisenatide, semaglutide or tirzepatide), including combination products (Supplementary Table 2).

### Covariates

We extracted primary care information on age, sex, ethnicity (supplemented with HES), practice geographical region, area-level deprivation quintile, primary care attendances over the prior six months, smoking, alcohol misuse, substance misuse, most recent HbA1c, glucose, estimated glomerular filtration rate (eGFR), weight and BMI, microvascular complications, cardiovascular and other physical comorbidities, and concomitant cardiovascular medications (full list in the Supplementary Methods).

### Statistical analysis

Full details of the statistical analyses are provided in the Supplementary Methods. For objective 1 (patient characteristics), variables were summarised at T2DM diagnosis in the incident T2DM cohort and at first GLP-1RA prescription among initiators, overall and by SMI status (no SMI, severe depression, bipolar disorder, other psychosis and schizophrenia), using medians and interquartile ranges (IQR) or counts and percentages.

For objective 2 (prescribing over time), we calculated the annual incidence of GLP-1RA prescribing per 1000 person-years in the full T2DM cohort, excluding individuals with GLP-1RA prescriptions prior to study start. We also calculated the point prevalence on 1 February each year in the full T2DM cohort, assuming a 90-day coverage window per prescription (Farmer et al., 2021). Both were reported for any GLP-1RA and for each formulation, overall and by SMI subtype, with SMI subtype recategorised each year using information available up to that time point.

For objective 3 (prescribing by SMI status), treatment lines were defined by the chronological sequence of dates on which one or more new antidiabetic classes were started. At treatment lines 2–5, among people intensifying at that timepoint (who had not previously received a GLP-1RA), we used logistic regression to estimate the odds of the new drug being a GLP-1RA rather than another antidiabetic, by SMI status (any SMI or each subtype, versus no SMI). We fitted marginal models; minimally adjusted models (age, sex, ethnicity, region, deprivation and calendar year with linear and quadratic terms); and fully adjusted models additionally including smoking, alcohol and substance misuse, primary care attendances, comorbidities, microvascular complications, concomitant medications, most recent BMI, HbA1c, eGFR and glucose, and biomarker recency, all measured at intensification. Confounders were selected using a directed acyclic graph (Supplementary Figure 1).

For objective 4 (treatment pathway), we described the antidiabetic classes initiated at each treatment line, prior antidiabetic class exposure among GLP-1RA initiators, and the time between treatment lines. We used Cox proportional hazards models with the same three covariate sets to compare time to intensification at lines 2–5 by SMI status.

For objective 5 (discontinuation and continuity of prescribing), we described GLP-1RA discontinuation (a gap of ≥60 days after the end of a prescription, assuming a 28-day supply) and restarting, as well as the proportion of days covered (PDC) as a measure of prescription coverage and proxy for adherence, with PDC ≥80% classified as high prescribing continuity over the specified period. Sensitivity analyses used a 90-day gap and dosage-derived supply (Supplementary Methods).

https://github.com/smwu/SMI_GLPMissing covariates were handled using multiple imputation by chained equations (25 imputed datasets per treatment line), assuming data were missing at random conditional on variables in the imputation model. Continuous variables were imputed using predictive mean matching, binary variables using logistic regression, and categorical variables using multinomial logistic regression. Estimates were pooled using Rubin’s rules, with complete-case analyses used as sensitivity analyses (Supplementary Methods). Analyses were conducted in R version 4.5.1 (R Core Team, 2025) using the mice package (Van Buuren & Groothuis-Oudshoorn, 2011).

### Lived experience involvement

This project was developed in consultation with people with lived experience of mental health diagnoses from the DATAMIND Lived Experience Advisory Group and the McPin Foundation. Both groups highlighted the adverse cardiometabolic effects of treatments for mental health, inequalities in access to cardiometabolic medication in people with mental illness, and the potential for these medications to impact mental health as research priorities that could have important clinical implications.

## Findings

### Patient characteristics

Of 2,102,329 adults with T2DM in the full cohort, 1,145,907 had incident T2DM (Supplementary Figure 2), of whom 320,751 (27.9%) had a prior diagnosis of SMI: 295,046 with severe depression, 6,833 with bipolar disorder, 7,982 with other psychosis and 10,890 with schizophrenia (Table 1). People with SMI were younger, more often female, more likely to live in the most deprived quintile, and had higher rates of smoking, alcohol, and substance misuse than the people without SMI. The schizophrenia and other psychosis subtypes had higher proportions of Black or Asian ethnicity. Glycaemic markers at diagnosis were similar across groups; weight and BMI were slightly higher in SMI. Cardiovascular disease, hypertension and cardiovascular medication use were more common in the non-SMI and depression groups (Supplementary Results).

**Table 1.** Sociodemographic and clinical characteristics of patients at the time of incident T2DM diagnosis, overall and by SMI subtype, between 2000 and 2025.

| Characteristic | Overall<br>n = 1,145,907 | No SMI<br>n = 825,156 | Any SMI<br>n = 320,751<sup>1</sup> | Depression<br>n = 295,046 | Bipolar disorder<br>n = 6,833 | Other psychosis<br>n = 7,982 | Schizophrenia<br>n = 10,890 |
|---|---|---|---|---|---|---|---|
| Age, years, median [IQR] | 61.0 [52.0, 71.0] | 62.0 [52.0, 72.0] | 59.0 [50.0, 68.0] | 59.0 [51.0, 68.0] | 56.0 [48.0, 65.0] | 56.0 [48.0, 67.0] | 54.0 [45.0, 63.0] |
| Year of T2DM diagnosis, median [IQR] | 2015 [2010, 2021] | 2015 [2010, 2020] | 2016 [2011, 2021] | 2016 [2011, 2021] | 2016 [2011, 2021] | 2016 [2011, 2021] | 2015 [2010, 2020] |
| **Sex, n (%)** |  |  |  |  |  |  |  |
| Female | 497,542 (43.4%) | 316,239 (38.3%) | 181,303 (56.5%) | 169,203 (57.3%) | 3,881 (56.8%) | 3,874 (48.5%) | 4,345 (39.9%) |
| Male | 648,365 (56.6%) | 508,917 (61.7%) | 139,448 (43.5%) | 125,843 (42.7%) | 2,952 (43.2%) | 4,108 (51.5%) | 6,545 (60.1%) |
| **Ethnicity, n (%)** |  |  |  |  |  |  |  |
| White | 857,225 (82.5%) | 597,989 (79.9%) | 259,236 (89.2%) | 240,795 (90.3%) | 5,447 (86.7%) | 5,651 (77.3%) | 7,343 (72.7%) |
| Black | 48,132 (4.6%) | 40,026 (5.3%) | 8,106 (2.8%) | 6,140 (2.3%) | 247 (3.9%) | 573 (7.8%) | 1,146 (11.4%) |
| Asian | 113,543 (10.9%) | 95,245 (12.7%) | 18,298 (6.3%) | 15,683 (5.9%) | 464 (7.4%) | 865 (11.8%) | 1,286 (12.7%) |
| Mixed | 8,808 (0.8%) | 6,686 (0.9%) | 2,122 (0.7%) | 1,733 (0.6%) | 70 (1.1%) | 113 (1.5%) | 206 (2.0%) |
| Other | 11,469 (1.1%) | 8,759 (1.2%) | 2,710 (0.9%) | 2,433 (0.9%) | 57 (0.9%) | 106 (1.5%) | 114 (1.1%) |
| Missing | *106,730 (9.3%)* | *76,451 (9.3%)* | *30,279 (9.4%)* | *28,262 (9.6%)* | *548 (8.0%)* | *674 (8.4%)* | *795 (7.3%)* |
| **Geographic region, n (%)** |  |  |  |  |  |  |  |
| East Midlands | 22,669 (2.0%) | 16,231 (2.0%) | 6,438 (2.0%) | 6,005 (2.0%) | 142 (2.1%) | 115 (1.4%) | 176 (1.6%) |
| East of England | 42,839 (3.7%) | 31,870 (3.9%) | 10,969 (3.4%) | 10,194 (3.5%) | 236 (3.5%) | 238 (3.0%) | 301 (2.8%) |
| London | 163,071 (14.2%) | 128,506 (15.6%) | 34,565 (10.8%) | 29,641 (10.0%) | 920 (13.5%) | 1,439 (18.0%) | 2,565 (23.6%) |
| North East | 42,395 (3.7%) | 27,970 (3.4%) | 14,425 (4.5%) | 13,472 (4.6%) | 280 (4.1%) | 324 (4.1%) | 349 (3.2%) |
| North West | 189,804 (16.6%) | 129,879 (15.7%) | 59,925 (18.7%) | 55,216 (18.7%) | 1,190 (17.4%) | 1,433 (18.0%) | 2,086 (19.2%) |
| Northern Ireland | 20,881 (1.8%) | 13,283 (1.6%) | 7,598 (2.4%) | 7,059 (2.4%) | 139 (2.0%) | 178 (2.2%) | 222 (2.0%) |
| Scotland | 89,957 (7.9%) | 63,145 (7.7%) | 26,812 (8.4%) | 24,906 (8.4%) | 552 (8.1%) | 619 (7.8%) | 735 (6.7%) |
| South East | 192,258 (16.8%) | 140,009 (17.0%) | 52,249 (16.3%) | 48,579 (16.5%) | 1,211 (17.7%) | 1,132 (14.2%) | 1,327 (12.2%) |
| South West | 107,040 (9.3%) | 76,699 (9.3%) | 30,341 (9.5%) | 28,299 (9.6%) | 589 (8.6%) | 687 (8.6%) | 766 (7.0%) |
| Wales | 64,683 (5.6%) | 44,110 (5.3%) | 20,573 (6.4%) | 19,209 (6.5%) | 414 (6.1%) | 457 (5.7%) | 493 (4.5%) |
| West Midlands | 179,145 (15.6%) | 130,886 (15.9%) | 48,259 (15.0%) | 44,455 (15.1%) | 988 (14.5%) | 1,181 (14.8%) | 1,635 (15.0%) |
| Yorkshire & The Humber | 31,165 (2.7%) | 22,568 (2.7%) | 8,597 (2.7%) | 8,011 (2.7%) | 172 (2.5%) | 179 (2.2%) | 235 (2.2%) |
| **2019 English IMD quintile, n (%)** |  |  |  |  |  |  |  |
| 1 (Least deprived) | 161,370 (17.1%) | 124,477 (18.1%) | 36,893 (14.2%) | 34,745 (14.6%) | 750 (13.4%) | 702 (10.7%) | 696 (7.5%) |
| 2 | 174,088 (18.4%) | 130,752 (19.0%) | 43,336 (16.7%) | 40,510 (17.0%) | 908 (16.3%) | 860 (13.1%) | 1,058 (11.5%) |
| 3 | 184,155 (19.5%) | 135,457 (19.7%) | 48,698 (18.8%) | 45,026 (18.9%) | 1,028 (18.4%) | 1,144 (17.4%) | 1,500 (16.3%) |
| 4 | 207,859 (22.0%) | 149,239 (21.7%) | 58,620 (22.6%) | 53,139 (22.3%) | 1,298 (23.3%) | 1,655 (25.2%) | 2,528 (27.4%) |
| 5 (Most deprived) | 218,559 (23.1%) | 146,629 (21.4%) | 71,930 (27.7%) | 64,687 (27.2%) | 1,593 (28.6%) | 2,207 (33.6%) | 3,443 (37.3%) |
| Missing | *199,876 (17%)* | *138,602 (17%)* | *61,274 (19%)* | *56,939 (19%)* | *1,256 (18%)* | *1,414 (18%)* | *1,665 (15%)* |
| **Prior substance use, n (%)** |  |  |  |  |  |  |  |
| Smoking | 824,214 (71.9%) | 574,456 (69.6%) | 249,758 (77.9%) | 229,248 (77.7%) | 5,493 (80.4%) | 6,227 (78.0%) | 8,790 (80.7%) |
| Alcohol misuse | 135,390 (11.8%) | 83,333 (10.1%) | 52,057 (16.2%) | 46,269 (15.7%) | 1,621 (23.7%) | 1,860 (23.3%) | 2,307 (21.2%) |
| Substance misuse | 19,947 (1.7%) | 5,320 (0.6%) | 14,627 (4.6%) | 10,928 (3.7%) | 832 (12.2%) | 1,126 (14.1%) | 1,741 (16.0%) |
| **Biomarkers, median [IQR]<sup>2</sup>** |  |  |  |  |  |  |  |
| HbA1c, mmol/mol | 53.0 [49.0, 66.0] | 53.0 [49.0, 67.0] | 52.0 [49.0, 63.0] | 52.0 [49.0, 63.0] | 53.0 [49.0, 64.0] | 53.0 [49.0, 66.0] | 53.0 [49.0, 66.0] |
| Missing | *286,374 (25%)* | *214,153 (26%)* | *72,221 (23%)* | *66,178 (22%)* | *1,497 (22%)* | *1,677 (21%)* | *2,869 (26%)* |
| Glucose, mmol/L | 8.0 [6.8, 11.8] | 8.0 [6.8, 12.0] | 7.9 [6.7, 11.3] | 7.9 [6.7, 11.2] | 8.1 [6.7, 11.6] | 8.0 [6.7, 11.9] | 8.2 [6.8, 12.3] |
| Missing | *418,026 (36%)* | *298,004 (36%)* | *120,022 (37%)* | *111,328 (38%)* | *2,283 (33%)* | *2,964 (37%)* | *3,447 (32%)* |
| eGFR, mL/min/1.73m² | 87.6 [72.2, 100.1] | 86.7 [71.4, 99.4] | 89.6 [74.1, 101.9] | 89.5 [74.0, 101.7] | 90.0 [73.3, 102.9] | 91.3 [75.3, 104.5] | 92.3 [76.6, 105.1] |
| Missing | *88,258 (7.7%)* | *67,718 (8.2%)* | *20,540 (6.4%)* | *18,685 (6.3%)* | *343 (5.0%)* | *564 (7.1%)* | *948 (8.7%)* |
| Weight, kg | 91.0 [78.0, 106.0] | 90.0 [77.2, 105.0] | 94.0 [80.0, 110.0] | 94.0 [80.0, 110.0] | 96.0 [82.5, 111.6] | 93.0 [79.0, 109.0] | 96.0 [82.0, 112.0] |
| Missing | *298,535 (26%)* | *223,936 (27%)* | *74,599 (23%)* | *70,952 (24%)* | *886 (13%)* | *1,341 (17%)* | *1,420 (13%)* |
| BMI, kg/m² | 31.9 [28.1, 36.7] | 31.3 [27.7, 35.8] | 33.5 [29.2, 38.7] | 33.5 [29.2, 38.7] | 34.0 [29.8, 39.1] | 33.0 [28.7, 38.0] | 33.3 [29.2, 38.4] |
| BMI ≥35 kg/m², n (%) | 278,660 (32.6%) | 175,017 (28.9%) | 103,643 (41.7%) | 94,445 (41.7%) | 2,672 (44.7%) | 2,636 (39.4%) | 3,890 (40.8%) |
| Missing | *291,043 (25%)* | *218,741 (27%)* | *72,302 (23%)* | *68,805 (23%)* | *857 (13%)* | *1,296 (16%)* | *1,344 (12%)* |
| **Comorbidities, n (%)** |  |  |  |  |  |  |  |
| Cardiovascular disease<sup>3</sup> | 272,246 (23.8%) | 191,949 (23.3%) | 80,297 (25.0%) | 75,822 (25.7%) | 1,212 (17.7%) | 1,686 (21.1%) | 1,577 (14.5%) |
| Myocardial infarction | 70,778 (6.2%) | 51,496 (6.2%) | 19,282 (6.0%) | 18,382 (6.2%) | 234 (3.4%) | 372 (4.7%) | 294 (2.7%) |
| Angina | 79,197 (6.9%) | 54,169 (6.6%) | 25,028 (7.8%) | 23,997 (8.1%) | 280 (4.1%) | 411 (5.1%) | 340 (3.1%) |
| Ischaemic heart disease<sup>4</sup> | 134,276 (11.7%) | 95,530 (11.6%) | 38,746 (12.1%) | 36,945 (12.5%) | 473 (6.9%) | 736 (9.2%) | 592 (5.4%) |
| Heart failure | 56,430 (4.9%) | 40,406 (4.9%) | 16,024 (5.0%) | 15,106 (5.1%) | 237 (3.5%) | 357 (4.5%) | 324 (3.0%) |
| Stroke | 61,537 (5.4%) | 41,484 (5.0%) | 20,053 (6.3%) | 18,881 (6.4%) | 316 (4.6%) | 449 (5.6%) | 407 (3.7%) |
| Peripheral arterial disease | 48,408 (4.2%) | 33,841 (4.1%) | 14,567 (4.5%) | 13,711 (4.6%) | 222 (3.2%) | 310 (3.9%) | 324 (3.0%) |
| Hypertension | 596,465 (52.1%) | 435,680 (52.8%) | 160,785 (50.1%) | 151,212 (51.3%) | 2,715 (39.7%) | 3,265 (40.9%) | 3,593 (33.0%) |
| Dyslipidaemia | 330,058 (28.8%) | 231,132 (28.0%) | 98,926 (30.8%) | 91,583 (31.0%) | 1,991 (29.1%) | 2,357 (29.5%) | 2,995 (27.5%) |
| Liver disease | 71,714 (6.3%) | 44,075 (5.3%) | 27,639 (8.6%) | 25,520 (8.6%) | 579 (8.5%) | 779 (9.8%) | 761 (7.0%) |
| Renal disease | 200,949 (17.5%) | 141,132 (17.1%) | 59,817 (18.6%) | 55,239 (18.7%) | 1,431 (20.9%) | 1,516 (19.0%) | 1,631 (15.0%) |
| **Prior medications prescribed, n (%)** |  |  |  |  |  |  |  |
| Anticoagulants | 111,934 (9.8%) | 79,642 (9.7%) | 32,292 (10.1%) | 30,465 (10.3%) | 532 (7.8%) | 725 (9.1%) | 570 (5.2%) |
| Antiplatelets | 340,285 (29.7%) | 238,607 (28.9%) | 101,678 (31.7%) | 95,969 (32.5%) | 1,635 (23.9%) | 2,063 (25.8%) | 2,011 (18.5%) |
| Antihypertensives | 721,344 (62.9%) | 505,778 (61.3%) | 215,566 (67.2%) | 202,207 (68.5%) | 3,957 (57.9%) | 4,558 (57.1%) | 4,844 (44.5%) |
| Lipid-regulating medications | 564,529 (49.3%) | 401,702 (48.7%) | 162,827 (50.8%) | 151,140 (51.2%) | 3,275 (47.9%) | 3,762 (47.1%) | 4,650 (42.7%) |
| **Healthcare utilisation and observation time, median [IQR]** |  |  |  |  |  |  |  |
| GP consultations in prior 6 months | 3.0 [0.0, 6.0] | 3.0 [0.0, 6.0] | 4.0 [0.0, 8.0] | 4.0 [0.0, 8.0] | 4.0 [0.0, 9.0] | 4.0 [0.0, 8.0] | 3.0 [0.0, 7.0] |
| Time registered before T2DM diagnosis, years | 17.1 [7.9, 28.8] | 17.1 [7.8, 29.0] | 17.3 [8.3, 28.5] | 17.7 [8.6, 28.9] | 12.6 [5.6, 23.0] | 13.7 [5.9, 25.0] | 11.8 [5.1, 21.7] |
| Follow-up after T2DM diagnosis, years | 5.7 [2.4, 10.3] | 5.8 [2.4, 10.5] | 5.5 [2.3, 9.9] | 5.5 [2.3, 9.9] | 5.1 [2.2, 9.2] | 4.7 [2.1, 9.0] | 5.5 [2.3, 9.8] |

<sup>1</sup> Any SMI includes depression, bipolar disorder, other psychosis, and schizophrenia.
<sup>2</sup> Biomarkers were defined based on the value closest to the index date, among test result values recorded between 2 years prior to 7 days after the index date.
<sup>3</sup> Cardiovascular disease (CVD) was defined as a composite of myocardial infarction, angina, coronary revascularisation procedures, ischaemic heart disease, stroke, cerebrovascular disease, heart failure, and peripheral arterial disease.
<sup>4</sup> Ischaemic heart disease (IHD) was defined as a composite of myocardial infarction, angina, coronary revascularisation procedures, and other general IHD terms.

### Characteristics of GLP-1RA initiators

Among the incident T2DM cohort, 102,041 (8.9%) people initiated GLP-1RAs: 14.5% for those with depression, 13.4% for bipolar disorder, 9.3% for other psychosis, 7.6% for schizophrenia and 6.9% for no SMI (Supplementary Table 3). 45,295 (44.4%) initiators had SMI, and the median year of initiation was 2022. Compared with the full incident cohort at diagnosis, initiators were younger (median 57.0 vs 61.0 years), more often White (88.8% vs 82.5%) and female (50.0% vs 43.4%), and had a substantially greater metabolic burden: median HbA1c 71.0 vs 53.0 mmol/mol, BMI 36.5 vs 31.9 kg/m², 59.1% vs 32.6% with class 2 or 3 obesity, and 38.4% with retinopathy. Hypertension, dyslipidaemia, liver and renal disease, and lipid-regulating and antihypertensive prescribing were also more common. Across SMI subtypes, over 50% of initiators had class 2 or 3 obesity (BMI ≥35), with the highest rates in depression (64.2%) and bipolar disorder (64.1%), while HbA1c levels were highest in bipolar disorder, other psychoses and schizophrenia. Initiators with schizophrenia were registered in less deprived areas than the corresponding T2DM populations, the reverse of the pattern in depression (Supplementary Results).

### Incidence and point prevalence over time

Incident GLP-1RA prescribing in the full T2DM cohort remained below 10 per 1000 person-years until 2018, dominated by exenatide and liraglutide. It then rose to 20.8 per 1000 in 2022 with the introduction of semaglutide and dulaglutide (Supplementary Figure 3). Rates fell in 2023 during national supply shortages (Supplementary Table 2) before increasing sharply to 33.9 in 2024 and 53.9 in 2025, driven by semaglutide and tirzepatide, which accounted for almost all new prescribing from 2024. When examining by SMI status, incidence was about 1.5 times higher in SMI than in no SMI, but this masked marked subtype heterogeneity. In 2025, incidence of any GLP-1RA was 74.6 per 1000 in depression (with tirzepatide alone reaching 92.1, indicating switching from other formulations), 66.7 in bipolar disorder, 46.4 in other psychosis and 31.6 in schizophrenia, compared with 44.3 in no SMI; the rate in schizophrenia was 2.4 times lower than in depression. Point prevalence in the full T2DM cohort increased more gradually, rising to 2.9% in 2013, plateauing until 2017, dipping in early 2024 during shortages and reaching 9.3% in 2025 (Supplementary Figure 4), again highest in depression and lowest in schizophrenia and psychosis.

### SMI status and GLP-1RA initiation at treatment intensification

The marginal odds of receiving a GLP-1RA rather than another antidiabetic at intensification were higher in the SMI group than in non-SMI at each treatment line, with odds ratios (ORs) ranging from 1.91 (95% confidence interval [CI] 1.85–1.96) at treatment line 2 to 1.25 (95% CI 1.19–1.30) at treatment line 5 (Figure 1). Adjustment for sociodemographic factors and calendar year attenuated these estimates (OR 1.37 [95% CI 1.32–1.41] and OR 1.05 [95% CI 1.01–1.10]), and further adjustment for diabetes progression factors attenuated them further (OR 1.20 [95% CI 1.16–1.24] and OR 0.99 [95% CI 0.94–1.04]). Overall SMI estimates were driven by the large depression subtype and concealed opposing patterns by subtype. After full adjustment, severe depression remained associated with higher odds of GLP-1RA initiation at treatment line 2 (OR 1.22 [95% CI 1.17–1.26]), treatment line 3 (OR 1.16 [95% CI 1.13–1.20]) and treatment line 4 (OR 1.06 [95% CI 1.03–1.10]), whereas schizophrenia was associated with lower odds at treatment line 2 (OR 0.74 [95% CI 0.61–0.88]) and line 3 (OR 0.72 [95% CI 0.62–0.84]), and other psychosis with lower odds at treatment line 3 (OR 0.82 [95% CI 0.69–0.96]). Odds ratio estimates for bipolar disorder were generally above 1 but not statistically significant. The complete-case analysis results were similar (Supplementary Figure 5).

**Figure 1.**
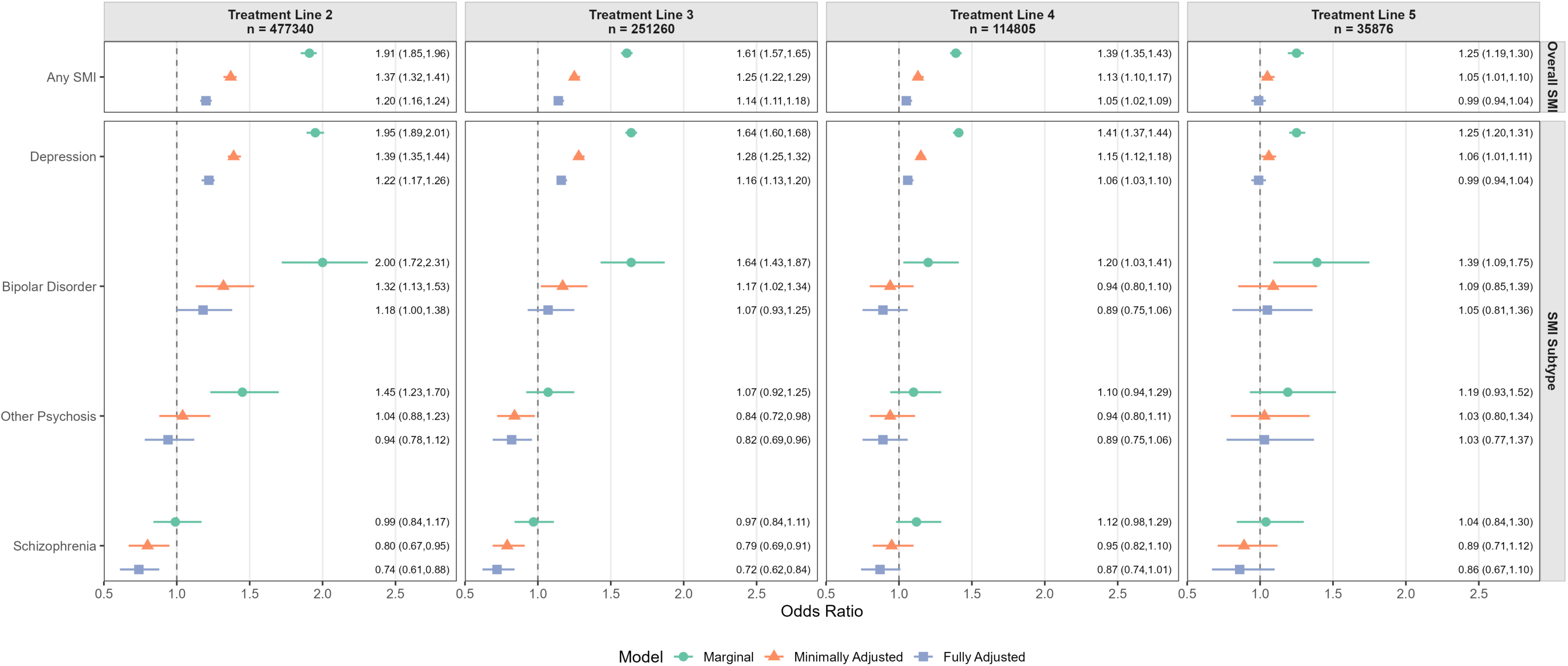
Association between severe mental illness and initiation of GLP-1RA therapy among individuals with type 2 diabetes. Points represent odds ratios from logistic regression models; horizontal lines represent 95% confidence intervals. The top panels show associations for any severe mental illness (SMI) versus no SMI. The bottom panels show associations for SMI subtypes, each compared with no SMI. Odds ratios greater than 1 indicate higher odds of GLP-1RA prescribing. Treatment line 2 represents second-line antidiabetic prescribing among those with prescriptions at that time; subsequent treatment lines are defined analogously. Minimally adjusted models included age, sex, region, area-level deprivation, ethnicity, and calendar year, with age centred and calendar year modelled using centred linear and quadratic terms. Fully adjusted models additionally included smoking status, alcohol misuse, substance misuse, GP attendances in the prior six months, most recent HbA1c, glucose, eGFR, and BMI, biomarker recency, hypertension, dyslipidaemia, liver disease, renal disease, cardiovascular disease, microvascular complications, and concomitant use of anticoagulants, antiplatelets, antihypertensives, and lipid-regulating medications.

### GLP-1RAs within the T2DM treatment pathway

GLP-1RAs were most often started as the third (27.9%) or fourth (28.5%) antidiabetic class, but 23.5% of initiators received them as first-or second-line treatment. This was slightly higher in those with SMI (26.5%; Table 2). People with SMI tended to start GLP-1RAs earlier in the pathway (55.1% before fourth line), except those with schizophrenia (Supplementary Table 4). Most initiators had prior exposure to metformin, sulfonylureas, DPP-4 inhibitors or SGLT-2 inhibitors; among 29,052 fourth-line initiators, 99.6% had received metformin, 64.9% DPP-4 inhibitors, 60.2% sulfonylureas and 55.2% SGLT-2 inhibitors (Supplementary Tables 4–5). Treatment intensification was more frequent and faster in SMI overall (25.5% vs 21.6% reached third-line or higher therapy; new treatment lines started 3–5 months earlier; Supplementary Table 4), but marginal hazard ratios (HR) of 1.14–1.21 for lines 2–5 fell to 1.04–1.09 after full adjustment (Supplementary Figure 6). Subtypes again diverged: fully adjusted hazards were highest in depression (HR 1.11 [95% CI 1.10–1.12] for intensification from line 1 to 2, to 1.05 [95% CI 1.03– 1.07] for line 4 to 5) and lowest in schizophrenia (0.81 [95% CI 0.78–0.83] to 0.90 [95% CI 0.82– 0.99]), with slower progression also in other psychosis for lines 1 to 2 (0.93, 95% CI 0.90–0.96) and 4 to 5 (0.87, 95% CI 0.79–0.97). Complete-case results were slightly attenuated but otherwise similar (Supplementary Figure 7).

**Table 2.** Column percentages of newly introduced antidiabetic classes by treatment line among patients with T2DM, for those with and without SMI Treatment lines are defined by the chronological sequence of dates on which one or more new antidiabetic drug classes were started. Treatment lines 6 and 7 are suppressed due to small counts.

| **Treatment Line** | **GLP-1RAs, n (%)** | **Metformin, n (%)** | **SGLT-2is, n (%)** | **DPP-4is, n (%)** | **Sulfonylureas, n (%)** | **Other, n (%)** | **Insulin, n (%)** |
|---|---|---|---|---|---|---|---|
| **Overall** |  |  |  |  |  |  |  |
| 1 | 4,789 (4.7%) | 792,624 (94.3%) | 26,082 (9.6%) | 11,852 (5.3%) | 67,116 (23.9%) | 1,981 (4.8%) | 19,305 (14.2%) |
| 2 | 19,138 (18.8%) | 44,247 (5.3%) | 115,918 (42.5%) | 110,380 (49.4%) | 153,707 (54.8%) | 17,391 (42.4%) | 27,647 (20.3%) |
| 3 | 28,446 (27.9%) | 3,554 (0.4%) | 71,162 (26.1%) | 79,023 (35.3%) | 41,410 (14.8%) | 13,563 (33.0%) | 32,086 (23.6%) |
| 4 | 29,052 (28.5%) | 372 (0.0%) | 40,119 (14.7%) | 18,271 (8.2%) | 13,708 (4.9%) | 5,193 (12.7%) | 32,340 (23.8%) |
| 5 | 16,196 (15.9%) | 61 (0.0%) | 14,359 (5.3%) | 3,347 (1.5%) | 3,587 (1.3%) | 2,053 (5.0%) | 18,035 (13.3%) |
| 6 | 4,048 (4.0%) | 9 (0.0%) | 4,496 (1.6%) | 671 (0.3%) | 695 (0.2%) | 725 (1.8%) | 5,939 (4.4%) |
| 7 | 372 (0.4%) | <5 | 704 (0.3%) | 75 (0.0%) | 58 (0.0%) | 138 (0.3%) | 702 (0.5%) |
| **No SMI** |  |  |  |  |  |  |  |
| 1 | 2,535 (4.1%) | 563,361 (94.2%) | 18,757 (9.9%) | 8,372 (5.3%) | 49,257 (24.8%) | 1,493 (5.0%) | 13,646 (15.4%) |
| 2 | 10,592 (17.3%) | 31,918 (5.3%) | 80,173 (42.2%) | 78,330 (49.5%) | 109,142 (54.9%) | 12,826 (43.3%) | 17,987 (20.3%) |
| 3 | 16,833 (27.4%) | 2,457 (0.4%) | 49,916 (26.3%) | 56,279 (35.6%) | 28,431 (14.3%) | 9,709 (32.8%) | 20,271 (22.9%) |
| 4 | 18,182 (29.6%) | 239 (0.0%) | 28,220 (14.8%) | 12,657 (8.0%) | 9,118 (4.6%) | 3,648 (12.3%) | 20,746 (23.4%) |
| 5 | 10,388 (16.9%) | 38 (0.0%) | 9,662 (5.1%) | 2,147 (1.4%) | 2,237 (1.1%) | 1,378 (4.7%) | 11,546 (13.0%) |
| 6 | 2,629 (4.3%) | <10 | 2,913 (1.5%) | 429 (0.3%) | 430 (0.2%) | 479 (1.6%) | 3,874 (4.4%) |
| 7 | 232 (0.4%) | <5 | 470 (0.2%) | 44 (0.0%) | 29 (0.0%) | 84 (0.3%) | 492 (0.6%) |
| **SMI** |  |  |  |  |  |  |  |
| 1 | 2,254 (5.5%) | 229,263 (94.4%) | 7,325 (8.9%) | 3,480 (5.3%) | 17,859 (21.9%) | 488 (4.3%) | 5,659 (11.9%) |
| 2 | 8,546 (21.0%) | 12,329 (5.1%) | 35,745 (43.2%) | 32,050 (49.0%) | 44,565 (54.6%) | 4,565 (39.9%) | 9,660 (20.3%) |
| 3 | 11,613 (28.6%) | 1,097 (0.5%) | 21,246 (25.7%) | 22,744 (34.8%) | 12,979 (15.9%) | 3,854 (33.7%) | 11,815 (24.9%) |
| 4 | 10,870 (26.7%) | 133 (0.1%) | 11,899 (14.4%) | 5,614 (8.6%) | 4,590 (5.6%) | 1,545 (13.5%) | 11,594 (24.4%) |
| 5 | 5,808 (14.3%) | 23 (0.0%) | 4,697 (5.7%) | 1,200 (1.8%) | 1,350 (1.7%) | 675 (5.9%) | 6,489 (13.7%) |
| 6 | 1,419 (3.5%) | <5 | 1,583 (1.9%) | 242 (0.4%) | 265 (0.3%) | 246 (2.2%) | 2,065 (4.3%) |
| 7 | 140 (0.3%) | <5 | 234 (0.3%) | 31 (0.0%) | 29 (0.0%) | 54 (0.5%) | 210 (0.4%) |

### Discontinuation of GLP-1Ras and continuity of prescribing after initiation

Over half of initiators discontinued (≥60-day gap) at least once during follow-up, rising from 53.6% in bipolar to 56.9% in schizophrenia (Figure 2). Fewer than a third discontinued by six months, but around 45% had discontinued by one year and around 60% by two years (63.0% in psychosis). Median time to first discontinuation among those who discontinued ranged from about six months in psychosis to eight months in depression, and 55.9–61.3% of those who discontinued later restarted (Supplementary Table 6). High prescribing continuity (PDC ≥80%) was uncommon overall, with the highest proportions in bipolar (46.6%) and schizophrenia (44.8%) and the lowest in the non-SMI group (38.2%) over available follow-up. By one year, fewer than half met this threshold in every group except bipolar and schizophrenia, in whom median PDC remained 79.0% and 73.7%, respectively, at two years. Sensitivity analyses using a 90-day gap or dosage-derived supply were generally consistent (Supplementary Table 7; Supplementary Results).

**Figure 2.**
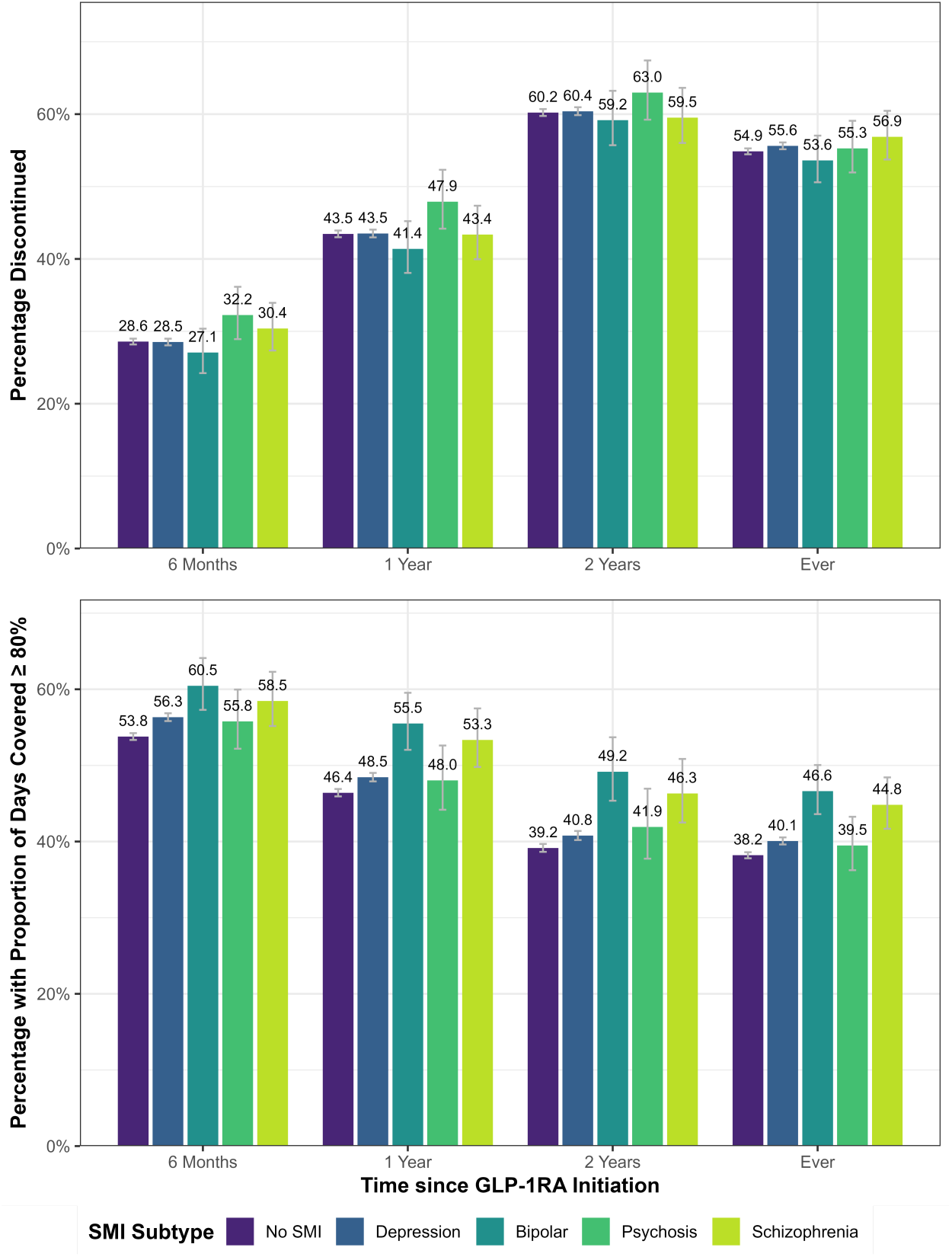
Treatment discontinuation and continuity of prescribing following GLP-1RA initiation, by SMI subtype. Percentage of patients who discontinued GLP-1RA treatment (top panel) and the proportion with a PDC ≥80% based on recorded prescribing (bottom panel), assessed at 6 months, 1 year, 2 years, and over the full available follow-up after GLP-1RA initiation. Results are shown for patients without SMI and for those with depression, bipolar disorder, psychosis, or schizophrenia. PDC reflects prescription coverage and does not confirm that medication was dispensed or taken. Bars show percentages, and error bars indicate 95% Wilson confidence intervals.

## Discussion

### Principal findings

In this 20-year study of over one million adults with incident T2DM in UK primary care, GLP-1RA prescribing was concentrated among those with substantial metabolic burden and rose steeply from 2019, accelerating in 2024–2025. Overall prescribing was higher among people with SMI, but this reflected prescribing in the large depression subtype and differences in demographic and clinical characteristics between the SMI and non-SMI groups. After adjustment, depression remained associated with modestly higher odds of GLP-1RA initiation and faster diabetes treatment intensification. In contrast schizophrenia and other psychosis were associated with lower odds of initiation and slower intensification. Almost a quarter of initiators received a GLP-1RA before the recommended third-or fourth-line treatment. Interruption of GLP-1RA prescribing and restarting were common across all groups, but continuity of prescribing was not lower in SMI and was highest in bipolar and schizophrenia.

### Comparison with other studies

The GLP-1RA initiators in our study had advanced diabetes and obesity, consistent with a previous UK description of GLP-1RA recipients as a selected population (Ulrich et al., 2026). They were disproportionately younger, female and White, and, among those with schizophrenia, from less deprived areas than the corresponding T2DM populations, echoing Swedish and US findings (Fors et al., 2025; Mittman et al., 2024). These comparisons were descriptive and assessed at different disease stages, but they support further investigation of inequalities in eligibility and uptake. The temporal pattern, with growth after the introduction of once-weekly formulations, a dip during the 2023–2024 shortages (Mahase, 2024) and rapid expansion of semaglutide and tirzepatide in 2025, means that the calendar period is tightly linked to formulation, indication and availability and must be accounted for in comparative effectiveness and safety studies. Prescribing is likely to rise further under the expanded eligibility in the 2026 NICE guideline (NICE, 2026).

Our overall SMI summary estimates did not reflect findings in our schizophrenia or psychosis subtypes. The overall excess of GLP1-RA prescribing in depression may reflect obesity severity, patient preference, or clinician recognition of weight-related need. Our definition required both a diagnosis and antidepressant treatment and therefore may have selected people with more consistently managed care, leaving the possibility of residual confounding by healthcare engagement. By contrast, the likelihood of receiving a GLP-1RA at the third treatment line was much lower in schizophrenia and in other psychosis, despite these subtypes having the highest HbA1c levels. This suggests a higher threshold for initiation in these SMI subtypes, consistent with meta-analytic evidence of poorer diabetes care for people with mental disorders (Wagner et al., 2026). Possible explanations include concerns about injectable treatment and adherence burden, more fragmented care, competing clinical priorities, and uncertainty about psychiatric effects of GLP1-RAs. Recent trial evidence shows metabolic benefit in antipsychotic-treated schizophrenia (Larsen et al., 2017; Ganeshalingam et al., 2025; Siskind et al., 2025). Evidence on psychiatric safety of GLP-1RAs is also increasingly reassuring (Pierret et al., 2025; Shapiro et al., 2025; Taipale et al., 2026), but people with SMI remain under-represented in that literature.

Although GLP-1RAs were typically introduced after metformin and several other classes, nearly a quarter of initiators received them at first or second line, for both those with and without SMI and particularly among those with depression, indicating real-world heterogeneity and some guideline discordance that should inform comparator selection in future observational studies. Among the incident T2DM cohort, around 45% of initiators had a 60-day prescription gap by one year and 60% by two years, similar to earlier CPRD estimates (Weiss et al., 2022). More than half restarted GLP1-RAs, consistent with US evidence that interruption and reinitiation are common (Rodriguez et al., 2025). Gaps may also reflect clinical stopping decisions or national shortages (Supplementary Tables 1 and 2); future studies should allow for repeated episodes and switching between formulations. We found no evidence that continuity of prescribing after GLP-1RA initiation was poorer in SMI. In fact, people with bipolar disorder or schizophrenia had the highest proportion with PDC ≥80% and the highest median PDC at two years, in keeping with meta-analytic evidence of higher diabetes medication adherence in schizophrenia (Gorczynski et al., 2017). The data captured prescriptions issued, not those dispensed or taken, so this higher prescription continuity may reflect structured contact with services or supported medication management. Initiators with schizophrenia, the subtype least likely to start, may be a selected subtype with better and more consistent access to healthcare in general.

### Strengths and limitations

Strengths include a large, broadly representative population with two decades of follow-up spanning major changes in GLP-1RA availability; examination of prescribing across the full treatment pathway; detailed covariate data enabling comparison of marginal and adjusted estimates; subtype-specific analyses; and consistent sensitivity analyses. Limitations include possible misclassification of diagnoses and prescriptions; possible underreporting of deaths in CPRD Gold practices after ONS coverage ended on 3 April 2024; absence of private and secondary care prescribing, dispensing and ingestion data; the fixed 28-day supply assumption (although dosage-derived results were similar); and lack of information on indications, contraindications, preferences and prescribing rationale. As such, adjusted associations should not be interpreted as causal effects of SMI or as direct measures of discrimination. Residual confounding (for example, by income, education or housing) and informative missingness in biomarkers are possible. Treatment-line models conditioned on reaching and intensifying at each line, and higher mortality in SMI, particularly schizophrenia, may have contributed to lower hazards of treatment intensification. Guidance, availability and indications changed substantially over the period, and the calendar year of 2025 was only partially observed.

## Clinical implications

GLP-1RA prescribing for T2DM in UK primary care has risen rapidly and is concentrated among people with high metabolic need. Higher crude prescribing in people with SMI overall concealed lower initiation and slower intensification in populations diagnosed with schizophrenia and psychosis: the subtypes at greatest cardiometabolic risk. Continuity of treatment after initiation was not worse in any SMI subtype. These findings support routine assessment of GLP-1RA eligibility within cardiometabolic care for people with SMI, particularly psychosis. Shared protocols between primary care, diabetes and mental health services may reduce fragmented responsibility, alongside supported decision-making and follow-up to help patients manage injections, adverse effects, supply interruptions and restarts. As eligibility expands, future monitoring should examine SMI subtypes separately at each stage of the pathway, from metabolic assessment and referral to treatment offer, initiation, dispensing and sustained use, to ensure that expansion reduces rather than reinforces physical health inequalities.

## Statements

### Data availability

Data underlying this study were accessed via CPRD under approved protocol no. 25_005368. Authors are not able to share the data directly; however, data can be accessed directly from CPRD following approval and licensing (see https://cprd.com/ for further details). The analytic code and materials supporting the findings are available in the online repository: https://github.com/smwu/SMI_GLP.

## Supporting information

RECORD-PE Checklist

## Acknowledgements

This study is based on data from the Clinical Practice Research Datalink (CPRD) obtained under licence from the UK Medicines and Healthcare products Regulatory Agency. Data are provided by patients and collected by the NHS as part of their care and support. The interpretation and conclusions contained in this study are those of the authors alone. The authors thank all of the patients and general practices contributing to the CPRD database.

## Author contributions

SMW, DPJO, and JFH formulated the research questions and designed the study. SMW and SP created code lists. SMW analysed the data. SMW and JFH wrote the first draft of the manuscript, and SP, NL, ARB, and DPJO critically reviewed the manuscript for important intellectual content. All authors approved the final version to be published and agree to be accountable for all aspects of the work in ensuring that questions related to the accuracy or integrity of any part of the work are appropriately investigated and resolved.

## Ethics and consent

CPRD has overarching research ethics approval from the Health Research Authority’s East Midlands-Derby Research Ethics Committee for observational research using anonymised patient data (reference: 21/EM/0265). The protocol for this study was approved through CPRD’s Research Data Governance process (protocol no. 25_005368). Individual patient consent was not required because the study used anonymised routinely collected data. Patients may opt out of their health records being used for research.

## Transparency Declaration

The corresponding author and manuscript guarantor affirm that the manuscript is an honest, accurate, and transparent account of the study being reported; that no important aspects of the study have been omitted; and that any discrepancies from the study as planned (and, if relevant, registered) have been explained.

