## Supplementary Materials for "Prescribing of glucagon-like peptide-1 receptor agonists for type 2 diabetes by severe mental illness status: a UK primary care cohort study, 2005-2025"

**Supplementary Methods**

#### Cohort definition

The study cohort comprised adults aged 18 to 100 who were actively registered with CPRD Gold or Aurum practices between 2005 and 2025, had at least 365 days of research quality primary care practice registration, and had type 2 diabetes mellitus (T2DM). T2DM diagnosis was defined as a recorded T2DM medical code or an antidiabetic prescription, confirmed by at least one laboratory test above threshold (glycated haemoglobin (HbA1c) ≥48 mmol/mol (6.5%) or fasting plasma glucose ≥7.0 mmol/L), based on NICE diagnostic guidelines and recommendations from previous literature on T2DM ascertainment in CPRD (NICE, 2025; Jeffery et al., 2024). HbA1c recordings not measured in % or mmol/mol, or with values equivalent to <4 or >250 mmol/mol, were excluded as unreliable. Values <20 were assumed to be measured as a %, and values ≥20 in mmol/mol. Where multiple measurements per patient per day were recorded, only the entry with the most reliable units was kept (see Processing of HbA1c and glucose laboratory measures sections below). We excluded all individuals initiating insulin within 6 months of the date of diagnosis as possible type 1 diabetes mellitus or very advanced diabetes, as well as individuals with T2DM or antidiabetic entries only present during periods of pregnancy as potential misclassification of gestational diabetes, following recommendations from previous research (Tate et al., 2017; Jeffery et al., 2024).

The date of diagnosis was defined as the earliest T2DM code or antidiabetic prescription date. The date of valid research-quality post-registration was defined as 365 days after the latest of registration start and the date at which data are considered adequate for research. Patients meeting eligibility criteria entered the study on the latest of 1 January 2005, the year individuals turned 18, the date of T2DM diagnosis, and the date of valid research-quality post-registration. Follow-up ended at the earliest of 31 May 2025, the year individuals turned 100, CPRD deregistration, the date of last collection, and ONS- or CPRD-recorded death. Those with missing information on biological sex were excluded (n=16). Cohort characteristics were calculated for all individuals with incident T2DM (i.e., diagnosed after the start of follow-up) using information up to and including the date of T2DM diagnosis, defined as baseline. Biomarker values for HbA1c, glucose, eGFR, weight, and BMI consisted of the most recent recorded value occurring within two years prior to baseline.

#### Processing of HbA1c laboratory measures

1. HbA1c values were allowed to be recorded in % (National Glycohemoglobin Standardization Program/Diabetes Control and Complications Trial (NGSP/DCCT)) or mmol/mol (International Federation of Clinical Chemistry and Laboratory Medicine (IFCC)). If no unit in % or mmol/mol was provided, values <20 were inferred to have unit %, and values ≥20 were inferred to have unit mmol/mol.
2. All values were converted to mmol/mol. For a given percentage $x$, the value in mmol/mol was given by: $\left( x - 2.152 \right) / 0.09148$.
3. Values outside of the analytical range of 4 to 250 mmol/mol (2.5% to 25%) were dropped as invalid measurements.
4. For each patient and day, only one measurement was kept. If there were multiple measurements, the measurements were ranked from most-to-least reliable unit in the following order: 1) recorded mmol/mol, 2) recorded %, 3) inferred mmol/mol, 4) inferred %. The most reliable unit was taken, using only the first entry if there were still multiple.

#### Processing of glucose laboratory measures

1. Glucose values were allowed to be recorded in mg/dL or mmol/L. If no unit in mg/dL or mmol/L was provided, values <35 were inferred to have unit mmol/L, and values ≥35 were inferred to have unit mg/dL.
2. All values were converted to mmol/L. For a given mg/dL value $x$, the equivalent value in mmol/mol was given by: $x \times0.0555$.
3. Values outside of the analytical range of 0.6 to 45 mmol/L were dropped as invalid measurements.
4. For each patient and day, only one measurement was kept. If there were multiple measurements, the measurements were ranked from most-to-least reliable unit in the following order: 1) recorded mmol/L, 2) recorded mg/dL, 3) inferred mmol/L, 4) inferred mg/dL. The most reliable unit was taken, using only the first entry if there were still multiple.

#### Severe mental illness definition

The exposure of severe mental illness (SMI) was defined as either: 1) ever receiving a diagnosis of schizophrenia, bipolar disorder, or other psychotic disorder; or 2) ever receiving a diagnosis of major depressive disorder and ever receiving a prescription of antidepressant medication. The date of SMI diagnosis was defined as the date of the first SMI diagnostic code, or the date of the first prescription of an appropriate drug treatment for that diagnosis if earlier (antipsychotic or mood stabiliser for schizophrenia, bipolar disorder, or other psychotic disorder; antidepressant for major depressive disorder). If a patient's record included more than one SMI subtype, the following classification hierarchy was used: 1) schizophrenia, 2) bipolar disorder, 3) other psychotic disorder, 4) major depressive disorder. For example, if a patient contributed follow-up from 2005 to 2025 and was diagnosed with a psychotic disorder in 2013, then diagnosed with schizophrenia in 2017, the patient would be classified as the other psychotic disorder subtype from 2013 to 2016, and as the schizophrenia subtype from 2017 to 2025. A CONSORT diagram for the SMI subpopulation is provided in Supplementary Figure 1. Code lists used to identify SMI diagnoses and treatments are publicly available at https://github.com/smwu/SMI_GLP.

#### GLP-1RA prescription definition

The outcome of GLP-1RA medication prescription was defined as a prescription, issued through primary care during the study period (2005-2025), of a medication with a primary active ingredient included under the class of GLP-1RA drugs, consisting of the following formulations: albiglutide, dulaglutide, exenatide, liraglutide, lixisenatide, semaglutide, or tirzepatide. Combination treatments including a GLP-1RA active ingredient were also included. To identify GLP-1RA medications, generic and common brand names were identified through reviewing national and international sources, then used to find relevant product codes in the CPRD code dictionaries and extract data from prescription records (Supplementary Table 2). Code lists used to identify GLP-1RA prescriptions are publicly available at <https://github.com/smwu/SMI_GLP>.

#### Covariates

Other variables of interest were ascertained from primary care records and included age, sex, ethnicity (supplemented with HES), geographic region, area-level relative deprivation, number of primary care attendances in the prior six months, time registered before T2DM diagnosis, follow-up after T2DM diagnosis, prior or current exposure to substances (smoking, alcohol misuse, substance misuse), most recent biomarker measurements (HbA1c, glucose, estimated glomerular filtration rate (eGFR) calculated from creatinine using the 2021 Chronic Kidney Disease Epidemiology Collaboration formula (Inker et al., 2021), weight, BMI), microvascular complications (retinopathy, nephropathy, neuropathy), physical health comorbidities (cardiovascular disease, myocardial infarction, angina, ischaemic heart disease, heart failure, stroke, peripheral arterial disease, hypertension, dyslipidaemia, liver disease, renal disease), and prior medications prescribed (anticoagulants, antiplatelets, antihypertensives, lipid-regulating medications). Ethnicity data for patients registered in England were supplemented with linked Hospital Episode Statistics data, where available. Geographic region referred to the location of the primary care practice at which a patient was registered. Area-level relative deprivation for patients registered in England referred to the quintile of the 2019 English Index of Multiple Deprivation, derived according to individuals' residential postcode or the practice postcode as a proxy.

#### Incidence and point prevalence

Annual incidence rates per 1000 person-years and point prevalence on 1 February of each calendar year were calculated in the full T2DM cohort that included individuals with either incident or pre-existing T2DM. Incidence and point prevalence were reported for any GLP-1RA and for each of the seven formulations, calculated overall and stratified by SMI group. Annual incidence rate was calculated as 1000 multiplied by the number of patients under follow-up and prescribed any GLP-1RA (or the relevant formulation for formulation-specific incidence) during each year, divided by the total follow-up time over all patients, with each patient contributing person-time until censoring or their first GLP-1RA prescription (or first prescription of the relevant formulation). To ensure identification of incident prescribing, patients with GLP-1RA prescriptions occurring prior to the start of follow-up were excluded (n = 25,280). To calculate incidence rates for each SMI group, all SMI diagnoses occurring before the end of each year were used to categorise individuals into mutually exclusive SMI exposure categories using the SMI hierarchy described above. Annual point prevalence was calculated as the percentage of patients who had at least one GLP-1RA prescription covering 1 February, among those still under follow-up on 1 February of the given year. We assumed a 90-day coverage window after each prescription, consisting of a 30-day base duration followed by a 60-day grace period; this has been shown to be comparable to more detailed dosage algorithms for antidiabetics using CPRD data (Farmer et al., 2021).

#### Treatment lines and regression models

Treatment lines were defined by the chronological sequence of dates on which one or more new antidiabetic drug classes were initiated. For the logistic regression models of GLP-1RA versus non-GLP-1RA prescription at treatment lines 2 through 5, we restricted each model to those who received treatment at that line and did not have prior initiation of GLP-1RAs. SMI status was defined as a binary exposure comparing SMI with no SMI (reference), then repeated using SMI as a categorical exposure with categories: no SMI (reference), depression, bipolar disorder, other psychosis, and schizophrenia. We ran three sets of models: 1) marginal; 2) minimally adjusted with sociodemographic covariates: age (centred), sex, ethnicity, geographic region, area-level relative deprivation, calendar year (modelled using centred linear and quadratic terms); and 3) fully adjusted with all covariates from the minimally adjusted model, as well as the following behavioural and clinical covariates measured at the time of treatment intensification: smoking status, alcohol misuse, substance misuse, GP attendances in the prior six months, comorbidities (cardiovascular disease, hypertension, dyslipidaemia, liver disease, and renal disease), microvascular complications (nephropathy, neuropathy, retinopathy), concomitant medications (anticoagulants, antiplatelets, antihypertensives, lipid-regulating medications), most recent cardiometabolic biomarker values (BMI, HbA1c, eGFR, glucose), and recency of biomarker measurements (≤6 months, >6 and ≤12 months, >12 months). These confounders were identified using a directed acyclic graph (Supplementary Figure 1).

For each treatment line (defined by the introduction of one or more antidiabetic classes), we obtained descriptive statistics of the number and percentage of individuals who initiated each of the following classes of antidiabetics: GLP-1RAs, metformin, SGLT-2is, DPP-4is, sulfonylureas, other (biguanides, thiazolidinediones, meglitinides, or alpha-glucosidase inhibitors), and insulin. Percentages were reported as both column percentages and row percentages. In addition, we reported the number and percentage of patients starting GLP-1RAs at each treatment line who had prior prescriptions of the other antidiabetic classes. We also examined the median number of days between T2DM diagnosis and each treatment line, as well as the median number of days between each treatment line. All of these descriptive statistics were calculated in the overall incident T2DM cohort, as well as stratified by the binary and categorical SMI exposure definitions. We modelled the association between time to treatment intensification (i.e., initiation of a new antidiabetic drug class) and SMI status (both binary and categorical versions) using Cox proportional hazards models for treatment lines 2 through 5. For each treatment line, analyses were restricted to individuals remaining under follow-up, with the index time point defined as the date of initiating the preceding treatment line. Marginal, minimally adjusted, and fully adjusted models were run using the same covariates as for the logistic regression models.

#### Missing data

Missing covariate values were addressed using multiple imputation by chained equations under a massing-at-random assumption conditional on observed variables in the imputation model. Separate imputation models were fitted for each treatment line to accommodate differences in populations at risk, missing-data patterns, and variable relationships across treatment stages. For each line, the imputation model included the SMI exposure and all covarites from the fully adjusted analysis. Logistic model imputations included the outcome, while Cox model imputations included the event indicator and Nelson-Aalen cumulative hazard estimator evaluated at each patient's event or censoring time. The same imputed datasets were used for the marginal and minimally adjusted analyses. Continuous variables were imputed using predictive mean matching, binary variables using logistic regression, and categorical variables using multinomial logistic regression. Twenty-five imputed datasets were generated per treatment line, and estimates were pooled using Rubin's rules. Multiple imputation was performed using the R package *mice* version 3.18.0 (Van Buuren & Groothuis-Oudshoorn, 2011). Complete-case analyses were performed as sensitivity analyses (Supplementary Figures 5 and 7).

#### Discontinuation, restarting, and continuity of prescribing

Treatment discontinuation was defined as a gap of at least 60 consecutive days after the end of the previous prescription (Ulrich et al., 2026). GLP-1RA prescriptions were assumed to cover 28 days. Time to first discontinuation was calculated from GLP-1RA initiation to the end of medication coverage preceding the first qualifying gap among patients who discontinued. The number of discontinuation episodes during follow-up was also calculated, and treatment restart was defined as a subsequent GLP-1RA prescription following a qualifying discontinuation. Continuity of prescribing was measured using the proportion of days covered (PDC) by prescriptions, calculated as the number of days covered by GLP-1RA treatment divided by the number of days in the prespecified observation window among those remaining under follow-up, allowing for carryover of oversupply within the observation window. PDC ≥80% was classified as high prescribing continuity over the specified period, as a proxy for adherence (Weiss et al., 2022). Discontinuation and continuity of prescribing were calculated at six months, one year, two years, and over all available follow-up. Continuous measures were summarised using medians and interquartile ranges, and categorical measures using counts and percentages. 95% Wilson confidence intervals were used to measure uncertainty in percentages. Sensitivity analyses were performed to assess robustness of results when defining the treatment discontinuation gap as at least 90 consecutive days, as well as calculating medication supply from dosage information available in CPRD (see Calculation of GLP-1RA medication supply below; Supplementary Table 7).

#### Calculation of GLP-1RA medication supply

We applied the following algorithm, adapted from Farmer et al. (2021) with clinician input, in sensitivity analyses where medication supply was explicitly calculated from dosage information available in CPRD:

1. Compute ‘quantity/qty’ divided by ‘daily_dose’, rounded to the nearest number of days, when both are available. If this is <1 or >90, set to NA.
2. If 1) is NA, use the value for ‘duration/numdays’. If ‘duration/numdays’ <1 or > 90, set to NA.
3. If 1) and 2) are NA, if ‘quantity’ >1 and <=90, use ‘quantity’ rounded to the nearest number of days. If not, set to NA.
4. If 1), 2), and 3) are all NA, assume 30-day prescription.

### **Supplementary Results**

#### Characteristics of the incident T2DM cohort by SMI subtype

Those with SMI were younger on average than those without SMI and had a higher proportion of females (Table 1). Ethnicity differed across SMI categories, with the schizophrenia and psychosis groups having a higher percentage identifying as Black or Asian (7.8%–12.7%), whereas the depression group was more predominantly White (90.3%). Geographically, a higher proportion (23.6%) of patients diagnosed with schizophrenia were registered in GP practices based in London, while this was only true for 10.0% of patients diagnosed with severe depression. In terms of area-level social deprivation, all SMI subtypes had a higher percentage of patients residing in the most deprived quintile (27.2%–37.3% in quintile 5) compared to those without SMI (21.4%). Prior or current exposure to smoking, alcohol misuse, and substance misuse were higher among the SMI groups compared to the non-SMI group. At the time of T2DM diagnosis, glycaemic biomarker values were similar across groups, while weight and BMI were slightly higher for the SMI groups. Among comorbidities, pre-existing cardiovascular diseases and hypertension were generally higher among the non-SMI and depression groups, while renal disease was more common in the bipolar disorder group. Prescriptions of anticoagulants, antiplatelets, antihypertensives, and lipid-regulating medications were generally higher in the non-SMI and depression groups.

#### Characteristics of GLP-1RA initiators

Compared to the full incident T2DM cohort, GLP-1RA initiators were younger (median age 57.0 at GLP-1RA initiation compared to median age 61.0 at time of T2DM diagnosis) and had higher percentages of White individuals (88.8% vs. 82.5%), females (50.0% vs. 43.4%), and prior or current exposure to smoking (79.7% vs. 71.9%), alcohol misuse (15.6% vs. 11.8%), and substance misuse (2.7% vs. 1.7%) (Supplementary Table 3). GLP-1RA initiators were registered at GP practices that were less concentrated in London (9.4% vs. 14.2%). Post-T2DM diagnosis follow-up also tended to be longer (median 10.0 vs. 5.7 years). Clinically, GLP-1RA initiators tended to have more metabolic burden, with higher glycaemic biomarker values at the time of initiation compared to baseline values among the full incident T2DM cohort (HbA1c 71.0 vs. 53.0 mmol/mol; glucose 9.8 vs. 8.0 mmol/L), as well as heavier weight (105.0 vs. 91.0 kg), higher BMI (36.5 vs. 31.9 kg/m²), and a higher proportion of patients with class 2 or 3 obesity (BMI $\geq$35 kg/ m²; 59.1% vs. 32.6%). GLP-1RA initiators also had higher rates of comorbid conditions, such as hypertension (63.2% vs. 52.1%), dyslipidaemia (38.4% vs. 28.8%), liver disease (15.6% vs. 6.3%), and renal disease (26.7% vs. 17.5%), as well as prior medication prescriptions, particularly for lipid-regulating medications (82.2% vs. 49.3%) and antihypertensives (77.3% vs. 62.9%), which were prescribed for over three quarters of GLP-1RA initiators.

GLP-1RA initiators were heterogeneous across SMI subtypes. Differentials for age, sex, ethnicity, geography, and prior exposure to substances were distributed similarly to those in the incident T2DM cohort, but this was not true for social deprivation, biomarker values, comorbidities, or prior medication prescriptions. Among patients with co-morbid schizophrenia, GLP-1RA initiators were more likely to reside in less deprived areas compared to the full T2DM cohort, while the converse trend was true for patients with co-morbid depression. Weight and BMI were elevated for all groups, with over 50% with class 2 or 3 obesity (BMI ≥35), particularly for the depression (64.2%) and bipolar (64.1%) groups, while median HbA1c values were higher among those with bipolar disorder, psychosis, or schizophrenia. Prior medication prescriptions were generally similar across all groups at the time of GLP-1RA initiation, although patients with schizophrenia had lower prior prescriptions for anticoagulants (7.5%), antiplatelets (23.9%), and antihypertensives (63.9%).

#### Incidence and point prevalence by formulation

Early incident prescribing (2005–2018) was largely composed of exenatide, liraglutide and, to a lesser extent, lixisenatide (Supplementary Figure 3). Incident prescribing generally increased from 2019 to 2022, coinciding with the introduction of semaglutide, with most initiations occurring for dulaglutide and semaglutide, as well as some liraglutide. Rates dropped in 2023, coinciding with drug shortage periods occurring in the latter half of the year (from July 2023 to December 2024 for liraglutide, exenatide, and dulaglutide; from September 2023 to June 2024 for semaglutide). In 2024 and 2025, incident prescribing increased sharply due to increases in prescribing of semaglutide and particularly the introduction of tirzepatide. There was very little incident prescribing of formulations other than semaglutide and tirzepatide from 2024. Prescribing of albiglutide was minimal throughout the study period up until its discontinuation in July 2018. General trends over time for the non-SMI group and the SMI subgroups were consistent with overall trends, with sharp increases in 2024 and 2025. Those with depression had the highest incidence rates, with tirzepatide incidence reaching 92.1 per 1000 person-years in 2025 and surpassing the overall GLP-1RA incidence rate (74.6), indicating switching to tirzepatide from other GLP-1RA formulations. Patients with bipolar disorder had the second highest incidence rates in 2025: 72.6 for tirzepatide and 66.7 for any GLP-1RA.

Point prevalence percentages calculated on 1 February of each year showed a more gradual ramp up of GLP-1RA prescribing, with prevalence rising to 2.9% in 2013, plateauing through 2017, then increasing steadily to 9.3% in 2025 (Supplementary Figure 4). Dulaglutide and semaglutide were the most prescribed formulations from 2022 to 2024, and semaglutide and tirzepatide were the most prescribed formulations in 2025. Prevalence dropped slightly in 2024, likely due to drug shortages that were not resolved until later in the year. Prescribing of liraglutide remained at around 1.5% from 2013 to 2023 but decreased in more recent years. Similar to incidence rates, point prevalence of GLP-1RA prescription was higher among those with SMI but differed substantially by subtype, with prevalence highest among those with depression and lowest among those with schizophrenia or psychosis.

#### Antidiabetic treatment pathway

The most commonly prescribed antidiabetics for each line were: metformin for line 1 (prescribed to 88.9% of those receiving first-line therapy); sulfonylureas for line 2 (prescribed to 32.1% of those receiving second-line therapy); DPP-4is (30.4%) and SGLT-2is (27.3%) for line 3; SGLT-2is (30.5%) for line 4; and insulin (32.8%) and GLP-1RAs (29.5%) for line 5 (Supplementary Table 4). These trends were consistent within the SMI group (Supplementary Table 4). Among those initiating GLP-1RAs, there was a high proportion with prior exposure to metformin, sulfonylureas, or SGLT-2 inhibitors (Supplementary Table 5). For example, among the 29,052 fourth-line GLP-1RA initiators, 99.6% had previous exposure to metformin, 64.9% to DPP-4is, 60.2% to sulfonylureas, and 55.2% to SGLT-2is.

Treatment intensification occurred more frequently, and more quickly, for patients with SMI (Supplementary Table 4). 25.5% of those with SMI received third-line or higher therapy (having failed first- and second-line therapy), whereas the same was true of only 21.6% of those without SMI. For second-line or higher treatments, patients with SMI started a new treatment line (i.e., incident prescription of a new antidiabetic) around 3–5 months earlier than their non-SMI counterparts (median 504–620 days for those with SMI; median 616–726 days for those without SMI). Median fourth-line treatment occurred 6.6 years (2421 days) and 7.9 years (2873 days) post-T2DM diagnosis for those with and without SMI, respectively. The association between SMI status and time to treatment intensification attenuated after adjustment for sociodemographic and clinical factors: marginal hazard ratios showed 14–21% higher hazards of intensification to treatment lines 2–5, which reduced to 4–9% after adjustment (Supplementary Figure 6).

#### Discontinuation and continuity of prescribing

Among patients still under follow-up, less than a third of patients discontinued GLP-1RA treatment by 6 months post-initiation, but this percentage increased to around 45% by 1 year post-initiation and around 60% by 2 years, with discontinuation most common among those with psychosis (63.0% by 2 years post-initiation) (Supplementary Table 6). Median time to first discontinuation ranged from around six months for those with psychosis to around eight months for those with depression. The majority (55.9%–61.3%) of patients who discontinued later restarted GLP-1RA treatment. PDC decreased steadily over time; however, patients with bipolar disorder or schizophrenia still maintained a median of 79.0% and 73.7%, respectively, of days covered by 2 years post-initiation. Results were consistent in sensitivity analyses using 90 consecutive days without prescription coverage as the discontinuation definition, as well as calculating medication supply using provided dosage information (Supplementary Table 7). Using a 90-day gap, discontinuation lowered from 55.2% to 49.1% at least once during follow-up, and from 28.6% to 23.8% by 6 months. Median time to first discontinuation extended to around 7.5-9.5 months, while the percentage who later restarted dropped to 48.1–54.9%. Using dosage-derived supply, the proportion with PDC ≥80% lowered from 39.1% to 38.7% over available follow-up, and from 55.0% to 50.1% at 6 months. By two years post-initiation, median PDC remained highest for bipolar (70.8%) and schizophrenia (62.1%), but the differential gap with other groups decreased.

### **Supplementary Tables**

#### Supplementary Table 1. UK historical NICE recommendations for GLP-1 receptor agonist prescribing, 2009–2025

| **Guidance period** | **Formulation(s) addressed** | **Treatment position** | **Key eligibility and continuation criteria** |
| --- | --- | --- | --- |
| **CG87**  **2009–2015** | Exenatide twice daily | Third therapy, added to metformin + sulfonylurea. | HbA1c ≥7.5%; BMI ≥35 kg/m² with weight-related problems, or BMI <35 kg/m² if insulin has occupational implications or weight loss would benefit an obesity-related comorbidity. Continue only with ≥1 percentage-point HbA1c reduction and ≥3% weight loss at 6 months. |
| **TA203**  **2010–2015** | Liraglutide 1.2 mg daily | Mainly third therapy in a triple regimen; restricted second-therapy use when conventional options are contraindicated or not tolerated. | Triple therapy uses the CG87 HbA1c/BMI criteria and stopping rule. Restricted dual therapy requires intolerance/contraindication to metformin or sulfonylurea and to both thiazolidinediones and DPP-4 inhibitors; continuation requires ≥1 percentage-point HbA1c reduction at 6 months. |
| **TA248**  **2012–2015** | Exenatide prolonged release once weekly | Same pathway position as TA203: mainly third therapy, with restricted second-therapy use. | Same principal initiation and continuation criteria as TA203. |
| **NG28**  **2015–March 2022** | Class-level recommendation | Later-line use after triple oral therapy failure. Prescribed as metformin + sulfonylurea + GLP-1RA, effectively replacing one oral drug. | BMI ≥35 kg/m² with obesity-related problems, or BMI <35 kg/m² when insulin has occupational implications or weight loss would benefit an obesity-related comorbidity. Continue only with ≥1 percentage-point HbA1c reduction and ≥3% weight loss at 6 months. |
| **NG28**  **March 2022–2025** | Class-level recommendation; evidence includes dulaglutide, exenatide, liraglutide, lixisenatide and semaglutide | Later-line use after ineffective, contraindicated or poorly tolerated triple oral therapy. Switch one drug for a GLP-1RA; the resulting regimen remains triple therapy. | BMI and continuation criteria as NG28. Removes the requirement to retain the specific metformin + sulfonylurea combination. GLP-1RA + insulin requires specialist support. |

**Sources:** NICE clinical guideline CG87, technology appraisals TA203 and TA248, and NICE guideline NG28 (original 2015 version and 2022 update). TA203 and TA248 supplemented CG87 rather than sequentially replacing it; all three were subsequently replaced by NG28 in 2015.

#### Supplementary Table 2. Summary of indication, administration and supply information for GLP-1RA medications authorised in the UK.

| Year Authorised | GLP-1RA Formulation | Brand Name | Indication | Mode of Administration | Notes |
| --- | --- | --- | --- | --- | --- |
| 2006 | Exenatide | Byetta | T2D | Twice daily injection | Shortage Jul 2023 to Mar 2024 discontinuation |
| 2009 | Liraglutide | Victoza | T2D | Once daily injection | Shortage Aug 2023 to Dec 2024 discontinuation |
| 2011 | Exenatide | Bydureon | T2D | Once weekly injection | Shortage Jul 2023 to Dec 2024 |
| 2013 | Lixisenatide | Lyxumia | T2D | Once daily injection | Shortage Jul 2023 to Dec 2023 discontinuation |
| 2014 | Albiglutide | Eperzan | T2D | Once weekly injection | Discontinued Jul 2018 |
| 2014 | Dulaglutide | Trulicity | T2D | Once weekly injection | Shortage Jul 2023 to Dec 2024 |
| 2014 | Insulin degludec + liraglutide | Xultophy | T2D | Once daily injection |  |
| 2015 | Liraglutide | Saxenda | WM | Once daily injection | Shortage Jul 2023 to Dec 2024 |
| 2017 | Insulin glargine + lixisenatide | Suliqua | T2D | Once daily injection |  |
| 2018 | Semaglutide | Ozempic | T2D | Once weekly injection | Shortage Jul 2023 to Jun 2024 |
| 2020 | Semaglutide | Rybelsus | T2D | Once daily oral tablet | Shortage Jul 2023 to Jan 2024 |
| 2022 | Semaglutide | Wegovy | WM | Once weekly injection | Shortage Sept 2023 to Dec 2024 |
| 2022 | Tirzepatide | Mounjaro | T2D | Once weekly injection |  |
| 2023 | Tirzepatide | Mounjaro | WM | Once weekly injection |  |
| 2024 | Semaglutide | Wegovy | CVD | Once weekly injection | Shortage Jul 2024 to Dec 2024 |
| Abbreviations: T2D = Type 2 diabetes, WM = weight management, CVD = cardiovascular disease. | | | | | |

#### Supplementary Table 3. Characteristics of individuals initiating GLP-1RA therapy among those with and without SMI

| Characteristic | Overall N = 102,041 | No SMI N = 56,746 | Any SMI N = 45,295^1^ | Depression N = 42,807 | Bipolar disorder N = 916 | Other psychosis N = 742 | Schizophrenia N = 830 |
| --- | --- | --- | --- | --- | --- | --- | --- |
| Age, years, median [IQR] | 57.0 [50.0, 65.0] | 59.0 [51.0, 66.0] | 56.0 [49.0, 63.0] | 56.0 [49.0, 63.0] | 54.0 [48.0, 61.0] | 55.0 [48.0, 61.0] | 53.0 [46.0, 60.0] |
| Year of GLP-1RA initiation, median [IQR] | 2022 [2018, 2024] | 2022 [2018, 2024] | 2021 [2018, 2024] | 2021 [2018, 2024] | 2022 [2019, 2024] | 2021 [2018, 2024] | 2021 [2018, 2023] |
| Sex, n (%) | | | | | | | |
| Female | 51,022 (50.0%) | 23,085 (40.7%) | 27,937 (61.7%) | 26,617 (62.2%) | 587 (64.1%) | 390 (52.6%) | 343 (41.3%) |
| Male | 51,019 (50.0%) | 33,661 (59.3%) | 17,358 (38.3%) | 16,190 (37.8%) | 329 (35.9%) | 352 (47.4%) | 487 (58.7%) |
| Ethnicity, n (%) | | | | | | | |
| White | 82,014 (88.8%) | 44,231 (86.1%) | 37,783 (92.0%) | 35,862 (92.6%) | 768 (90.2%) | 567 (82.1%) | 586 (76.0%) |
| Black | 2,893 (3.1%) | 2,037 (4.0%) | 856 (2.1%) | 706 (1.8%) | 21 (2.5%) | 45 (6.5%) | 84 (10.9%) |
| Asian | 5,928 (6.4%) | 4,113 (8.0%) | 1,815 (4.4%) | 1,628 (4.2%) | 42 (4.9%) | 59 (8.5%) | 86 (11.2%) |
| Mixed | 720 (0.8%) | 429 (0.8%) | 291 (0.7%) | 265 (0.7%) | 10 (1.2%) | 10 (1.4%) | 6 (0.8%) |
| Other | 847 (0.9%) | 532 (1.0%) | 315 (0.8%) | 286 (0.7%) | 10 (1.2%) | 10 (1.4%) | 9 (1.2%) |
| Missing | *9,639 (9.4%)* | *5,404 (9.5%)* | *4,235 (9.3%)* | *4,060 (9.5%)* | *65 (7.1%)* | *51 (6.9%)* | *59 (7.1%)* |
| Geographic region, n (%) | | | | | | | |
| East Midlands | 1,449 (1.4%) | 792 (1.4%) | 657 (1.5%) | 626 (1.5%) | 15 (1.6%) | 9 (1.2%) | 7 (0.8%) |
| East of England | 2,388 (2.3%) | 1,364 (2.4%) | 1,024 (2.3%) | 976 (2.3%) | 19 (2.1%) | 8 (1.1%) | 21 (2.5%) |
| London | 9,612 (9.4%) | 6,068 (10.7%) | 3,544 (7.8%) | 3,171 (7.4%) | 87 (9.5%) | 120 (16.2%) | 166 (20.0%) |
| North East | 3,206 (3.1%) | 1,613 (2.8%) | 1,593 (3.5%) | 1,517 (3.5%) | 27 (2.9%) | 26 (3.5%) | 23 (2.8%) |
| North West | 18,947 (18.6%) | 9,832 (17.3%) | 9,115 (20.1%) | 8,607 (20.1%) | 183 (20.0%) | 142 (19.1%) | 183 (22.0%) |
| Northern Ireland | 2,888 (2.8%) | 1,427 (2.5%) | 1,461 (3.2%) | 1,405 (3.3%) | 14 (1.5%) | 13 (1.8%) | 29 (3.5%) |
| Scotland | 8,445 (8.3%) | 4,752 (8.4%) | 3,693 (8.2%) | 3,506 (8.2%) | 73 (8.0%) | 61 (8.2%) | 53 (6.4%) |
| South East | 20,281 (19.9%) | 11,413 (20.1%) | 8,868 (19.6%) | 8,414 (19.7%) | 197 (21.5%) | 133 (17.9%) | 124 (14.9%) |
| South West | 9,903 (9.7%) | 5,418 (9.5%) | 4,485 (9.9%) | 4,284 (10.0%) | 67 (7.3%) | 76 (10.2%) | 58 (7.0%) |
| Wales | 6,545 (6.4%) | 3,455 (6.1%) | 3,090 (6.8%) | 2,945 (6.9%) | 59 (6.4%) | 47 (6.3%) | 39 (4.7%) |
| West Midlands | 16,219 (15.9%) | 9,347 (16.5%) | 6,872 (15.2%) | 6,510 (15.2%) | 157 (17.1%) | 95 (12.8%) | 110 (13.3%) |
| Yorkshire & The Humber | 2,158 (2.1%) | 1,265 (2.2%) | 893 (2.0%) | 846 (2.0%) | 18 (2.0%) | 12 (1.6%) | 17 (2.0%) |
| 2019 English IMD quintile, n (%) | | | | | | | |
| 1 (Least deprived) | 12,897 (15.6%) | 8,082 (17.4%) | 4,815 (13.2%) | 4,592 (13.4%) | 95 (12.5%) | 69 (11.3%) | 59 (8.4%) |
| 2 | 14,657 (17.7%) | 8,734 (18.9%) | 5,923 (16.3%) | 5,640 (16.4%) | 118 (15.5%) | 75 (12.3%) | 90 (12.8%) |
| 3 | 15,638 (18.9%) | 8,879 (19.2%) | 6,759 (18.6%) | 6,377 (18.6%) | 149 (19.6%) | 99 (16.2%) | 134 (19.1%) |
| 4 | 18,174 (22.0%) | 9,946 (21.5%) | 8,228 (22.6%) | 7,736 (22.5%) | 164 (21.6%) | 151 (24.7%) | 177 (25.2%) |
| 5 (Most deprived) | 21,393 (25.8%) | 10,685 (23.1%) | 10,708 (29.4%) | 10,012 (29.1%) | 235 (30.9%) | 218 (35.6%) | 243 (34.6%) |
| Missing | *19,282 (19%)* | *10,420 (18%)* | *8,862 (20%)* | *8,450 (20%)* | *155 (17%)* | *130 (18%)* | *127 (15%)* |
| Prior substance use, n (%) | | | | | | | |
| Smoking | 81,309 (79.7%) | 44,042 (77.6%) | 37,267 (82.3%) | 35,139 (82.1%) | 770 (84.1%) | 634 (85.4%) | 724 (87.2%) |
| Alcohol misuse | 15,875 (15.6%) | 7,997 (14.1%) | 7,878 (17.4%) | 7,219 (16.9%) | 241 (26.3%) | 210 (28.3%) | 208 (25.1%) |
| Substance misuse | 2,746 (2.7%) | 564 (1.0%) | 2,182 (4.8%) | 1,707 (4.0%) | 152 (16.6%) | 136 (18.3%) | 187 (22.5%) |
| Biomarkers, median [IQR]^2^ | | | | | | | |
| HbA1c, mmol/mol | 71.0 [60.0, 85.0] | 71.0 [61.0, 85.0] | 71.0 [59.0, 86.0] | 71.0 [59.0, 85.0] | 72.0 [58.0, 89.0] | 75.0 [61.0, 91.0] | 76.0 [62.0, 91.0] |
| Missing | *549 (0.5%)* | *345 (0.6%)* | *204 (0.5%)* | *183 (0.4%)* | *10 (1.1%)* | *<5* | *8 (1.0%)* |
| Glucose, mmol/L | 9.8 [7.4, 13.7] | 9.8 [7.4, 13.7] | 9.7 [7.3, 13.8] | 9.7 [7.3, 13.7] | 10.1 [7.2, 14.8] | 9.8 [7.2, 14.3] | 10.7 [7.6, 15.0] |
| Missing | *20,610 (20%)* | *11,305 (20%)* | *9,305 (21%)* | *8,898 (21%)* | *170 (19%)* | *133 (18%)* | *104 (13%)* |
| eGFR, mL/min/1.73m² | 97.2 [82.1, 106.0] | 96.2 [80.6, 105.2] | 98.4 [84.0, 107.0] | 98.3 [83.9, 106.9] | 99.2 [82.8, 108.6] | 99.4 [84.8, 108.0] | 100.7 [86.5, 109.5] |
| Missing | *526 (0.5%)* | *365 (0.6%)* | *161 (0.4%)* | *145 (0.3%)* | *8 (0.9%)* | *<5* | *5 (0.6%)* |
| Weight, kg | 105.0 [91.0, 120.9] | 104.1 [90.8, 120.0] | 105.0 [91.6, 122.0] | 105.0 [91.5, 122.0] | 104.6 [92.0, 120.7] | 107.0 [92.0, 125.0] | 106.0 [93.0, 121.0] |
| Missing | *1,024 (1.0%)* | *613 (1.1%)* | *411 (0.9%)* | *391 (0.9%)* | *8 (0.9%)* | *<5* | *9 (1.1%)* |
| BMI, kg/m² | 36.5 [32.3, 41.7] | 35.7 [31.8, 40.8] | 37.4 [33.1, 42.8] | 37.4 [33.1, 42.8] | 37.6 [33.0, 42.8] | 37.4 [32.7, 43.0] | 36.4 [32.5, 41.5] |
| BMI $\boldsymbol{\geq}$35 kg/m², n (%) | 59,694 (59.1%) | 30,897 (55.0%) | 28,797 (64.1%) | 27,259 (64.2%) | 583 (64.1%) | 471 (63.6%) | 484 (59.0%) |
| Missing | *1,013 (1.0%)* | *620 (1.1%)* | *393 (0.9%)* | *375 (0.9%)* | *7 (0.8%)* | *<5* | *9 (1.1%)* |
| Comorbidities, n (%) | | | | | | | |
| Cardiovascular disease^3^ | 24,772 (24.3%) | 13,763 (24.3%) | 11,009 (24.3%) | 10,483 (24.5%) | 179 (19.5%) | 188 (25.3%) | 159 (19.2%) |
| Myocardial infarction | 5,491 (5.4%) | 3,268 (5.8%) | 2,223 (4.9%) | 2,147 (5.0%) | 34 (3.7%) | 25 (3.4%) | 17 (2.0%) |
| Angina | 5,645 (5.5%) | 2,925 (5.2%) | 2,720 (6.0%) | 2,629 (6.1%) | 34 (3.7%) | 33 (4.4%) | 24 (2.9%) |
| Ischaemic heart disease^4^ | 10,816 (10.6%) | 6,093 (10.7%) | 4,723 (10.4%) | 4,519 (10.6%) | 71 (7.8%) | 81 (10.9%) | 52 (6.3%) |
| Heart failure | 5,090 (5.0%) | 2,994 (5.3%) | 2,096 (4.6%) | 1,992 (4.7%) | 36 (3.9%) | 35 (4.7%) | 33 (4.0%) |
| Stroke | 5,089 (5.0%) | 2,530 (4.5%) | 2,559 (5.6%) | 2,456 (5.7%) | 37 (4.0%) | 36 (4.9%) | 30 (3.6%) |
| Peripheral arterial disease | 5,991 (5.9%) | 3,417 (6.0%) | 2,574 (5.7%) | 2,431 (5.7%) | 31 (3.4%) | 59 (8.0%) | 53 (6.4%) |
| Hypertension | 64,459 (63.2%) | 36,866 (65.0%) | 27,593 (60.9%) | 26,280 (61.4%) | 489 (53.4%) | 425 (57.3%) | 399 (48.1%) |
| Dyslipidaemia | 39,188 (38.4%) | 21,480 (37.9%) | 17,708 (39.1%) | 16,713 (39.0%) | 360 (39.3%) | 289 (38.9%) | 346 (41.7%) |
| Liver disease | 15,878 (15.6%) | 7,790 (13.7%) | 8,088 (17.9%) | 7,620 (17.8%) | 159 (17.4%) | 160 (21.6%) | 149 (18.0%) |
| Renal disease | 27,233 (26.7%) | 15,394 (27.1%) | 11,839 (26.1%) | 11,129 (26.0%) | 283 (30.9%) | 206 (27.8%) | 221 (26.6%) |
| Diabetic complications, n (%) | | | | | | | |
| Retinopathy | 39,144 (38.4%) | 22,682 (40.0%) | 16,462 (36.3%) | 15,503 (36.2%) | 334 (36.5%) | 284 (38.3%) | 341 (41.1%) |
| Nephropathy | 1,054 (1.0%) | 677 (1.2%) | 377 (0.8%) | 355 (0.8%) | 10 (1.1%) | <5 | 9 (1.1%) |
| Neuropathy | 2,306 (2.3%) | 1,162 (2.0%) | 1,144 (2.5%) | 1,090 (2.5%) | 16 (1.7%) | 23 (3.1%) | 15 (1.8%) |
| Prior medications prescribed, n (%) | | | | | | | |
| Anticoagulants | 11,901 (11.7%) | 6,648 (11.7%) | 5,253 (11.6%) | 4,998 (11.7%) | 104 (11.4%) | 89 (12.0%) | 62 (7.5%) |
| Antiplatelets | 34,256 (33.6%) | 18,660 (32.9%) | 15,596 (34.4%) | 14,866 (34.7%) | 282 (30.8%) | 250 (33.7%) | 198 (23.9%) |
| Antihypertensives | 78,831 (77.3%) | 43,326 (76.4%) | 35,505 (78.4%) | 33,746 (78.8%) | 664 (72.5%) | 565 (76.1%) | 530 (63.9%) |
| Lipid-regulating medications | 83,888 (82.2%) | 46,943 (82.7%) | 36,945 (81.6%) | 34,888 (81.5%) | 735 (80.2%) | 632 (85.2%) | 690 (83.1%) |
| Healthcare utilisation and observation time, median [IQR] | | | | | | | |
| GP consultations in prior 6 months | 4.0 [0.0, 8.0] | 4.0 [0.0, 7.0] | 5.0 [0.0, 9.0] | 5.0 [0.0, 9.0] | 5.5 [0.0, 11.0] | 5.0 [0.0, 11.0] | 5.0 [0.0, 10.0] |
| Time registered before T2DM diagnosis, years | 15.5 [7.4, 25.5] | 15.6 [7.4, 25.9] | 15.4 [7.4, 25.2] | 15.6 [7.5, 25.4] | 12.2 [5.6, 20.9] | 12.5 [5.9, 21.5] | 10.7 [4.8, 20.7] |
| Follow-up after T2DM diagnosis, years | 10.0 [5.7, 14.5] | 10.3 [6.0, 14.7] | 9.6 [5.3, 14.1] | 9.6 [5.3, 14.1] | 8.6 [5.2, 13.1] | 8.8 [5.0, 13.0] | 10.0 [6.5, 13.9] |
| All characteristics are calculated at the time of first GLP-1RA prescription.  ^1^ Any SMI includes depression, bipolar disorder, other psychosis, and schizophrenia.  ^2^ Biomarkers were defined based on the value closest to the index date, among test result values recorded between 2 years prior to 7 days after the index date.  ^3^ Cardiovascular disease (CVD) was defined as a composite of myocardial infarction, angina, coronary revascularisation procedures, ischaemic heart disease, stroke, cerebrovascular disease, heart failure, and peripheral arterial disease.  ^4^ Ischaemic heart disease (IHD) was defined as a composite of myocardial infarction, angina, coronary revascularisation procedures, and other general IHD terms. | | | | | | | |

#### Supplementary Table 4. Antidiabetic treatment line progression and newly introduced antidiabetic classes at each treatment line among patients with incident T2DM, overall and by SMI status and subtype

Treatment lines are defined by the chronological sequence of dates on which one or more new antidiabetic drug classes were started. % of previous line = percentage of patients at the previous line who progressed to this line; % of cohort = percentage of the incident T2DM cohort in that stratum reaching this line. Column % = percentage of all initiations of the drug class that occurred at this treatment line; row % = percentage of patients starting a new class at this line who started the drug class. Small counts are suppressed for disclosure reasons.

| **Treatment line** | **Patients starting a new class, n** | **% of previous line** | **% of cohort** | **Median days since previous line** | **Median days since T2DM diagnosis** | **GLP-1RAs, n (column %; row %)** | **Metformin, n (column %; row %)** | **SGLT-2is, n (column %; row %)** | **DPP-4is, n (column %; row %)** | **Sulfonylureas, n (column %; row %)** | **Other, n (column %; row %)** | **Insulin, n (column %; row %)** |
| --- | --- | --- | --- | --- | --- | --- | --- | --- | --- | --- | --- | --- |
| **Overall (incident T2DM cohort, N = 1,145,907)** | | | | | | | | | | | | |
| 1 | 891,390 | 77.8 | 77.8 | 11 | 11.0 | 4,789 (4.7; 0.5) | 792,624 (94.3; 88.9) | 26,082 (9.6; 2.9) | 11,852 (5.3; 1.3) | 67,116 (23.9; 7.5) | 1,981 (4.8; 0.2) | 19,305 (14.2; 2.2) |
| 2 | 478,718 | 53.7 | 41.8 | 657 | 993.0 | 19,138 (18.8; 4.0) | 44,247 (5.3; 9.2) | 115,918 (42.5; 24.2) | 110,380 (49.4; 23.1) | 153,707 (54.8; 32.1) | 17,391 (42.4; 3.6) | 27,647 (20.3; 5.8) |
| 3 | 260,305 | 54.4 | 22.7 | 638 | 1932.0 | 28,446 (27.9; 10.9) | 3,554 (0.4; 1.4) | 71,162 (26.1; 27.3) | 79,023 (35.3; 30.4) | 41,410 (14.8; 15.9) | 13,563 (33.0; 5.2) | 32,086 (23.6; 12.3) |
| 4 | 131,625 | 50.6 | 11.5 | 596 | 2719.0 | 29,052 (28.5; 22.1) | 372 (0.0; 0.3) | 40,119 (14.7; 30.5) | 18,271 (8.2; 13.9) | 13,708 (4.9; 10.4) | 5,193 (12.7; 3.9) | 32,340 (23.8; 24.6) |
| 5 | 54,993 | 41.8 | 4.8 | 582 | 3380.0 | 16,196 (15.9; 29.5) | 61 (0.0; 0.1) | 14,359 (5.3; 26.1) | 3,347 (1.5; 6.1) | 3,587 (1.3; 6.5) | 2,053 (5.0; 3.7) | 18,035 (13.3; 32.8) |
| 6 | 16,289 | 29.6 | 1.4 | 636 | 3962.0 | 4,048 (4.0; 24.9) | 9 (0.0; 0.1) | 4,496 (1.6; 27.6) | 671 (0.3; 4.1) | 695 (0.2; 4.3) | 725 (1.8; 4.5) | 5,939 (4.4; 36.5) |
| 7 | 2,050 | 12.6 | 0.2 | 684 | 4455.5 | 372 (0.4; 18.1) | <5 | 704 (0.3; 34.3) | 75 (0.0; 3.7) | 58 (0.0; 2.8) | 138 (0.3; 6.7) | 702 (0.5; 34.2) |
| **No SMI (incident T2DM cohort, N = 825,156)** | | | | | | | | | | | | |
| 1 | 634,151 | 76.9 | 76.9 | 12 | 12 | 2,535 (4.1; 0.4) | 563,361 (94.2; 88.8) | 18,757 (9.9; 3.0) | 8,372 (5.3; 1.3) | 49,257 (24.8; 7.8) | 1,493 (5.0; 0.2) | 13,646 (15.4; 2.2) |
| 2 | 334,941 | 52.8 | 40.6 | 688 | 1045 | 10,592 (17.3; 3.2) | 31,918 (5.3; 9.5) | 80,173 (42.2; 23.9) | 78,330 (49.5; 23.4) | 109,142 (54.9; 32.6) | 12,826 (43.3; 3.8) | 17,987 (20.3; 5.4) |
| 3 | 178,579 | 53.3 | 21.6 | 691 | 2044 | 16,833 (27.4; 9.4) | 2,457 (0.4; 1.4) | 49,916 (26.3; 28.0) | 56,279 (35.6; 31.5) | 28,431 (14.3; 15.9) | 9,709 (32.8; 5.4) | 20,271 (22.9; 11.4) |
| 4 | 88,129 | 49.4 | 10.7 | 646 | 2873 | 18,182 (29.6; 20.6) | 239 (0.0; 0.3) | 28,220 (14.8; 32.0) | 12,657 (8.0; 14.4) | 9,118 (4.6; 10.3) | 3,648 (12.3; 4.1) | 20,746 (23.4; 23.5) |
| 5 | 35,732 | 40.5 | 4.3 | 616 | 3554 | 10,388 (16.9; 29.1) | 38 (0.0; 0.1) | 9,662 (5.1; 27.0) | 2,147 (1.4; 6.0) | 2,237 (1.1; 6.3) | 1,378 (4.7; 3.9) | 11,546 (13.0; 32.3) |
| 6 | 10,559 | 29.6 | 1.3 | 654 | 4143 | 2,629 (4.3; 24.9) | <10 | 2,913 (1.5; 27.6) | 429 (0.3; 4.1) | 430 (0.2; 4.1) | 479 (1.6; 4.5) | 3,874 (4.4; 36.7) |
| 7 | 1,351 | 12.8 | 0.2 | 726 | 4600 | 232 (0.4; 17.2) | <5 | 470 (0.2; 34.8) | 44 (0.0; 3.3) | 29 (0.0; 2.1) | 84 (0.3; 6.2) | 492 (0.6; 36.4) |
| **Any SMI (incident T2DM cohort, N = 320,751)** | | | | | | | | | | | | |
| 1 | 257,239 | 80.2 | 80.2 | 10.0 | 10 | 2,254 (5.5; 0.9) | 229,263 (94.4; 89.1) | 7,325 (8.9; 2.8) | 3,480 (5.3; 1.4) | 17,859 (21.9; 6.9) | 488 (4.3; 0.2) | 5,659 (11.9; 2.2) |
| 2 | 143,777 | 55.9 | 44.8 | 588.0 | 882 | 8,546 (21.0; 5.9) | 12,329 (5.1; 8.6) | 35,745 (43.2; 24.9) | 32,050 (49.0; 22.3) | 44,565 (54.6; 31.0) | 4,565 (39.9; 3.2) | 9,660 (20.3; 6.7) |
| 3 | 81,726 | 56.8 | 25.5 | 533.0 | 1709 | 11,613 (28.6; 14.2) | 1,097 (0.5; 1.3) | 21,246 (25.7; 26.0) | 22,744 (34.8; 27.8) | 12,979 (15.9; 15.9) | 3,854 (33.7; 4.7) | 11,815 (24.9; 14.5) |
| 4 | 43,496 | 53.2 | 13.6 | 504.0 | 2421 | 10,870 (26.7; 25.0) | 133 (0.1; 0.3) | 11,899 (14.4; 27.4) | 5,614 (8.6; 12.9) | 4,590 (5.6; 10.6) | 1,545 (13.5; 3.6) | 11,594 (24.4; 26.7) |
| 5 | 19,261 | 44.3 | 6.0 | 524.0 | 3052 | 5,808 (14.3; 30.2) | 23 (0.0; 0.1) | 4,697 (5.7; 24.4) | 1,200 (1.8; 6.2) | 1,350 (1.7; 7.0) | 675 (5.9; 3.5) | 6,489 (13.7; 33.7) |
| 6 | 5,730 | 29.7 | 1.8 | 601.5 | 3636 | 1,419 (3.5; 24.8) | <5 | 1,583 (1.9; 27.6) | 242 (0.4; 4.2) | 265 (0.3; 4.6) | 246 (2.2; 4.3) | 2,065 (4.3; 36.0) |
| 7 | 699 | 12.2 | 0.2 | 620.0 | 4142 | 140 (0.3; 20.0) | <5 | 234 (0.3; 33.5) | 31 (0.0; 4.4) | 29 (0.0; 4.1) | 54 (0.5; 7.7) | 210 (0.4; 30.0) |
| **Depression (incident T2DM cohort, N = 295,046)** | | | | | | | | | | | | |
| 1 | 236,575 | 80.2 | 80.2 | 10 | 10.0 | 2,146 (5.6; 0.9) | 210,688 (94.4; 89.1) | 6,893 (8.9; 2.9) | 3,170 (5.2; 1.3) | 16,264 (21.6; 6.9) | 438 (4.1; 0.2) | 5,101 (11.5; 2.2) |
| 2 | 133,341 | 56.4 | 45.2 | 591 | 889.0 | 8,087 (21.0; 6.1) | 11,272 (5.1; 8.5) | 33,513 (43.2; 25.1) | 29,700 (49.0; 22.3) | 41,013 (54.5; 30.8) | 4,218 (39.8; 3.2) | 8,963 (20.2; 6.7) |
| 3 | 76,193 | 57.1 | 25.8 | 532 | 1712.0 | 11,016 (28.7; 14.5) | 1,006 (0.5; 1.3) | 19,857 (25.6; 26.1) | 21,072 (34.8; 27.7) | 12,075 (16.1; 15.8) | 3,586 (33.8; 4.7) | 11,056 (24.9; 14.5) |
| 4 | 40,798 | 53.5 | 13.8 | 502 | 2419.0 | 10,266 (26.7; 25.2) | 123 (0.1; 0.3) | 11,090 (14.3; 27.2) | 5,281 (8.7; 12.9) | 4,321 (5.7; 10.6) | 1,451 (13.7; 3.6) | 10,879 (24.5; 26.7) |
| 5 | 18,195 | 44.6 | 6.2 | 525 | 3057.0 | 5,458 (14.2; 30.0) | 23 (0.0; 0.1) | 4,443 (5.7; 24.4) | 1,127 (1.9; 6.2) | 1,276 (1.7; 7.0) | 635 (6.0; 3.5) | 6,173 (13.9; 33.9) |
| 6 | 5,413 | 29.7 | 1.8 | 609 | 3644.0 | 1,341 (3.5; 24.8) | <5 | 1,499 (1.9; 27.7) | 228 (0.4; 4.2) | 254 (0.3; 4.7) | 231 (2.2; 4.3) | 1,948 (4.4; 36.0) |
| 7 | 662 | 12.2 | 0.2 | 631 | 4178.5 | 131 (0.3; 19.8) | <5 | 227 (0.3; 34.3) | 29 (0.0; 4.4) | 27 (0.0; 4.1) | 50 (0.5; 7.6) | 197 (0.4; 29.8) |
| **Bipolar disorder (incident T2DM cohort, N = 6,833)** | | | | | | | | | | | | |
| 1 | 5,536 | 81.0 | 81.0 | 9.0 | 9.0 | 56 (7.2; 1.0) | 4,906 (93.9; 88.6) | 114 (7.6; 2.1) | 106 (7.8; 1.9) | 445 (25.6; 8.0) | 14 (7.3; 0.3) | 145 (14.8; 2.6) |
| 2 | 2,935 | 53.0 | 43.0 | 546.0 | 792.0 | 180 (23.0; 6.1) | 292 (5.6; 9.9) | 654 (43.6; 22.3) | 661 (48.9; 22.5) | 946 (54.4; 32.2) | 77 (39.9; 2.6) | 207 (21.2; 7.1) |
| 3 | 1,587 | 54.1 | 23.2 | 493.0 | 1612.0 | 228 (29.2; 14.4) | 20 (0.4; 1.3) | 400 (26.7; 25.2) | 459 (33.9; 28.9) | 244 (14.0; 15.4) | 59 (30.6; 3.7) | 235 (24.1; 14.8) |
| 4 | 824 | 51.9 | 12.1 | 504.5 | 2310.5 | 176 (22.5; 21.4) | 5 (0.1; 0.6) | 227 (15.1; 27.5) | 104 (7.7; 12.6) | 81 (4.7; 9.8) | 25 (13.0; 3.0) | 252 (25.8; 30.6) |
| 5 | 338 | 41.0 | 4.9 | 435.5 | 2940.5 | 118 (15.1; 34.9) | <5 | 78 (5.2; 23.1) | 19 (1.4; 5.6) | 24 (1.4; 7.1) | 11 (5.7; 3.3) | 99 (10.1; 29.3) |
| 6 | 89 | 26.3 | 1.3 | 412.0 | 3480.0 | 21 (2.7; 23.6) | <5 | 25 (1.7; 28.1) | <5 | <5 | <5 | 35 (3.6; 39.3) |
| 7 | 11 | 12.4 | 0.2 | 1093.0 | 3492.0 | <5 | <5 | <5 | <5 | <5 | <5 | <5 |
| **Other psychosis (incident T2DM cohort, N = 7,982)** | | | | | | | | | | | | |
| 1 | 6,362 | 79.7 | 79.7 | 8.0 | 8.0 | 30 (4.5; 0.5) | 5,687 (93.9; 89.4) | 158 (9.4; 2.5) | 101 (7.3; 1.6) | 491 (24.9; 7.7) | 16 (6.5; 0.3) | 169 (16.7; 2.7) |
| 2 | 3,223 | 50.7 | 40.4 | 530.0 | 753.0 | 147 (21.8; 4.6) | 333 (5.5; 10.3) | 734 (43.5; 22.8) | 690 (49.6; 21.4) | 1,102 (55.9; 34.2) | 82 (33.3; 2.5) | 219 (21.6; 6.8) |
| 3 | 1,692 | 52.5 | 21.2 | 520.5 | 1574.5 | 170 (25.3; 10.0) | 36 (0.6; 2.1) | 442 (26.2; 26.1) | 478 (34.3; 28.3) | 262 (13.3; 15.5) | 94 (38.2; 5.6) | 255 (25.1; 15.1) |
| 4 | 839 | 49.6 | 10.5 | 566.0 | 2413.0 | 192 (28.5; 22.9) | <5 | 252 (14.9; 30.0) | 96 (6.9; 11.4) | 81 (4.1; 9.7) | 34 (13.8; 4.1) | 223 (22.0; 26.6) |
| 5 | 325 | 38.7 | 4.1 | 450.0 | 2889.0 | 105 (15.6; 32.3) | <5 | 68 (4.0; 20.9) | 22 (1.6; 6.8) | 28 (1.4; 8.6) | 15 (6.1; 4.6) | 102 (10.1; 31.4) |
| 6 | 110 | 33.8 | 1.4 | 449.0 | 3478.0 | 25 (3.7; 22.7) | <5 | 31 (1.8; 28.2) | <5 | 6 (0.3; 5.5) | <10 | 41 (4.0; 37.3) |
| 7 | 12 | 10.9 | 0.2 | 473.5 | 3340.5 | <5 | <5 | <5 | <5 | <5 | <5 | 5 (0.5; 41.7) |
| **Schizophrenia (incident T2DM cohort, N = 10,890)** | | | | | | | | | | | | |
| 1 | 8,766 | 80.5 | 80.5 | 8.0 | 8.0 | 22 (2.9; 0.3) | 7,982 (94.4; 91.1) | 160 (7.9; 1.8) | 103 (5.1; 1.2) | 659 (24.4; 7.5) | 20 (5.3; 0.2) | 244 (20.6; 2.8) |
| 2 | 4,278 | 48.8 | 39.3 | 572.0 | 822.5 | 132 (17.6; 3.1) | 432 (5.1; 10.1) | 844 (41.8; 19.7) | 999 (49.7; 23.4) | 1,504 (55.8; 35.2) | 188 (49.6; 4.4) | 271 (22.9; 6.3) |
| 3 | 2,254 | 52.7 | 20.7 | 595.5 | 1786.0 | 199 (26.5; 8.8) | 35 (0.4; 1.6) | 547 (27.1; 24.3) | 735 (36.6; 32.6) | 398 (14.8; 17.7) | 115 (30.3; 5.1) | 269 (22.7; 11.9) |
| 4 | 1,035 | 45.9 | 9.5 | 566.0 | 2584.0 | 236 (31.5; 22.8) | <5 | 330 (16.3; 31.9) | 133 (6.6; 12.9) | 107 (4.0; 10.3) | 35 (9.2; 3.4) | 240 (20.3; 23.2) |
| 5 | 403 | 38.9 | 3.7 | 557.0 | 3152.0 | 127 (16.9; 31.5) | <5 | 108 (5.3; 26.8) | 32 (1.6; 7.9) | 22 (0.8; 5.5) | 14 (3.7; 3.5) | 115 (9.7; 28.5) |
| 6 | 118 | 29.3 | 1.1 | 577.5 | 3583.5 | 32 (4.3; 27.1) | <5 | 28 (1.4; 23.7) | 6 (0.3; 5.1) | 5 (0.2; 4.2) | 6 (1.6; 5.1) | 41 (3.5; 34.7) |
| 7 | 14 | 11.9 | 0.1 | 308.5 | 3793.0 | <5 | <5 | <5 | <5 | <5 | <5 | <5 |

#### Supplementary Table 5. Prior antidiabetic prescribing among patients with incident T2DM initiating GLP-1RA therapy, overall and by SMI status and subtype

Percentages for a given row are calculated using the number of patients starting GLP-1RA for the treatment line as the denominator.

| **Treatment Line** | **Patients starting GLP-1RA, n** | **Median days since T2DM diagnosis** | **Previous Metformin, n (%)** | **Previous SGLT-2is, n (%)** | **Previous DPP-4is, n (%)** | **Previous Sulfonylureas, n (%)** | **Previous Other, n (%)** | **Previous Insulin, n (%)** |
| --- | --- | --- | --- | --- | --- | --- | --- | --- |
| **Overall** | | | | | | | | |
| 1 | 4789 | 11.0 | 0 (0.0%) | 0 (0.0%) | 0 (0.0%) | 0 (0.0%) | 0 (0.0%) | 0 (0.0%) |
| 2 | 19,138 | 961.0 | 18,049 (94.3%) | 819 (4.3%) | 240 (1.3%) | 953 (5.0%) | 32 (0.2%) | 623 (3.3%) |
| 3 | 28,446 | 1733.0 | 28,081 (98.7%) | 12,707 (44.7%) | 6,554 (23.0%) | 8,039 (28.3%) | 646 (2.3%) | 2,828 (9.9%) |
| 4 | 29,052 | 2521.0 | 28,942 (99.6%) | 16,031 (55.2%) | 19,285 (66.4%) | 17,555 (60.4%) | 2,553 (8.8%) | 4,003 (13.8%) |
| 5 | 16,196 | 3351.5 | 16,172 (99.9%) | 11,869 (73.3%) | 14,323 (88.4%) | 14,744 (91.0%) | 3,495 (21.6%) | 4,518 (27.9%) |
| 6 | 4,048 | 4033.5 | 4,045 (99.9%) | 3,509 (86.7%) | 3,945 (97.5%) | 3,959 (97.8%) | 2,237 (55.3%) | 2,587 (63.9%) |
| 7 | 372 | 4722.5 | 372 (100.0%) | 372 (100.0%) | 372 (100.0%) | 372 (100.0%) | 372 (100.0%) | 372 (100.0%) |
| **No SMI** | | | | | | | | |
| 1 | 2535 | 12.0 | 0 (0.0%) | 0 (0.0%) | 0 (0.0%) | 0 (0.0%) | 0 (0.0%) | 0 (0.0%) |
| 2 | 105,92 | 1052.0 | 9,977 (94.2%) | 475 (4.5%) | 136 (1.3%) | 566 (5.3%) | 23 (0.2%) | 385 (3.6%) |
| 3 | 16,833 | 1868.0 | 16,615 (98.7%) | 7,520 (44.7%) | 3,930 (23.3%) | 4,746 (28.2%) | 431 (2.6%) | 1,619 (9.6%) |
| 4 | 18,182 | 2670.0 | 18,118 (99.6%) | 9,956 (54.8%) | 12,227 (67.2%) | 11,009 (60.5%) | 1,694 (9.3%) | 2,334 (12.8%) |
| 5 | 10,388 | 3562.5 | 10,378 (99.9%) | 7,682 (74.0%) | 9,272 (89.3%) | 9,474 (91.2%) | 2,349 (22.6%) | 2,617 (25.2%) |
| 6 | 2,629 | 4239.0 | 2,626 (99.9%) | 2,276 (86.6%) | 2,563 (97.5%) | 2,576 (98.0%) | 1,553 (59.1%) | 1,584 (60.3%) |
| 7 | 232 | 4858.5 | 232 (100.0%) | 232 (100.0%) | 232 (100.0%) | 232 (100.0%) | 232 (100.0%) | 232 (100.0%) |
| **SMI** | | | | | | | | |
| 1 | 2,254 | 9.5 | 0 (0.0%) | 0 (0.0%) | 0 (0.0%) | 0 (0.0%) | 0 (0.0%) | 0 (0.0%) |
| 2 | 8,546 | 868.0 | 8,072 (94.5%) | 344 (4.0%) | 104 (1.2%) | 387 (4.5%) | 9 (0.1%) | 238 (2.8%) |
| 3 | 11,613 | 1552.0 | 11,466 (98.7%) | 5,187 (44.7%) | 2,624 (22.6%) | 3,293 (28.4%) | 215 (1.9%) | 1,209 (10.4%) |
| 4 | 10,870 | 2288.0 | 10,824 (99.6%) | 6,075 (55.9%) | 7,058 (64.9%) | 6,546 (60.2%) | 859 (7.9%) | 1,669 (15.4%) |
| 5 | 5,808 | 2995.5 | 5,794 (99.8%) | 4,187 (72.1%) | 5,051 (87.0%) | 5,270 (90.7%) | 1,146 (19.7%) | 1,901 (32.7%) |
| 6 | 1,419 | 3692.0 | 1,419 (100.0%) | 1,233 (86.9%) | 1,382 (97.4%) | 1,383 (97.5%) | 684 (48.2%) | 1,003 (70.7%) |
| 7 | 140 | 4429.0 | 140 (100.0%) | 140 (100.0%) | 140 (100.0%) | 140 (100.0%) | 140 (100.0%) | 140 (100.0%) |
| **Depression** | | | | | | | | |
| 1 | 2146 | 12.0 | 0 (0.0%) | 0 (0.0%) | 0 (0.0%) | 0 (0.0%) | 0 (0.0%) | 0 (0.0%) |
| 2 | 8087 | 863.0 | 7,638 (94.4%) | 327 (4.0%) | 94 (1.2%) | 354 (4.4%) | 8 (0.1%) | 218 (2.7%) |
| 3 | 11016 | 1555.5 | 10,875 (98.7%) | 4,955 (45.0%) | 2,491 (22.6%) | 3,099 (28.1%) | 203 (1.8%) | 1,119 (10.2%) |
| 4 | 10266 | 2282.0 | 10,221 (99.6%) | 5,752 (56.0%) | 6,669 (65.0%) | 6,160 (60.0%) | 810 (7.9%) | 1,574 (15.3%) |
| 5 | 5458 | 2997.0 | 5,445 (99.8%) | 3,937 (72.1%) | 4,748 (87.0%) | 4,949 (90.7%) | 1,090 (20.0%) | 1,768 (32.4%) |
| 6 | 1341 | 3706.0 | 1,341 (100.0%) | 1,164 (86.8%) | 1,305 (97.3%) | 1,310 (97.7%) | 649 (48.4%) | 945 (70.5%) |
| 7 | 131 | 4413.0 | 131 (100.0%) | 131 (100.0%) | 131 (100.0%) | 131 (100.0%) | 131 (100.0%) | 131 (100.0%) |
| **Bipolar Disorder** | | | | | | | | |
| 1 | 56 | 0.0 | 0 (0.0%) | 0 (0.0%) | 0 (0.0%) | 0 (0.0%) | 0 (0.0%) | 0 (0.0%) |
| 2 | 180 | 1055.0 | 173 (96.1%) | 5 (2.8%) | <5 | 13 (7.2%) | 0 (0.0%) | <5 |
| 3 | 228 | 1376.5 | 225 (98.7%) | 97 (42.5%) | 55 (24.1%) | 74 (32.5%) | 6 (2.6%) | 22 (9.6%) |
| 4 | 176 | 2095.0 | 176 (100.0%) | 95 (54.0%) | 126 (71.6%) | 104 (59.1%) | 7 (4.0%) | 33 (18.8%) |
| 5 | 118 | 2812.5 | 117 (99.2%) | 78 (66.1%) | 100 (84.7%) | 107 (90.7%) | 22 (18.6%) | 52 (44.1%) |
| 6 | 21 | 3235.0 | 21 (100.0%) | 19 (90.5%) | 21 (100.0%) | 20 (95.2%) | 7 (33.3%) | 17 (81.0%) |
| 7 | <5 | 4518.0 | <5 | <5 | <5 | <5 | <5 | <5 |
| **Other Psychosis** | | | | | | | | |
| 1 | 30 | 6.0 | 0 (0.0%) | 0 (0.0%) | 0 (0.0%) | 0 (0.0%) | 0 (0.0%) | 0 (0.0%) |
| 2 | 147 | 885.0 | 138 (93.9%) | 5 (3.4%) | 6 (4.1%) | 7 (4.8%) | 0 (0.0%) | 9 (6.1%) |
| 3 | 170 | 1298.0 | 168 (98.8%) | 58 (34.1%) | 34 (20.0%) | 62 (36.5%) | <5 | 35 (20.6%) |
| 4 | 192 | 2358.5 | 192 (100.0%) | 113 (58.9%) | 116 (60.4%) | 122 (63.5%) | 19 (9.9%) | 27 (14.1%) |
| 5 | 105 | 2705.0 | 105 (100.0%) | 76 (72.4%) | 89 (84.8%) | 98 (93.3%) | 17 (16.2%) | 39 (37.1%) |
| 6 | 25 | 2679.0 | 25 (100.0%) | 21 (84.0%) | 24 (96.0%) | 23 (92.0%) | 16 (64.0%) | 16 (64.0%) |
| 7 | <5 | 3868.5 | <5 | <5 | <5 | <5 | <5 | <5 |
| **Schizophrenia** | | | | | | | | |
| 1 | 22 | 26.0 | 0 (0.0%) | 0 (0.0%) | 0 (0.0%) | 0 (0.0%) | 0 (0.0%) | 0 (0.0%) |
| 2 | 132 | 978.0 | 123 (93.2%) | 7 (5.3%) | <5 | 13 (9.8%) | <5 | 9 (6.8%) |
| 3 | 199 | 1535.0 | 198 (99.5%) | 77 (38.7%) | 44 (22.1%) | 58 (29.1%) | <5 | 33 (16.6%) |
| 4 | 236 | 2533.5 | 235 (99.6%) | 115 (48.7%) | 147 (62.3%) | 160 (67.8%) | 23 (9.7%) | 35 (14.8%) |
| 5 | 127 | 3134.0 | 127 (100.0%) | 96 (75.6%) | 114 (89.8%) | 116 (91.3%) | 17 (13.4%) | 42 (33.1%) |
| 6 | 32 | 3899.0 | 32 (100.0%) | 29 (90.6%) | 32 (100.0%) | 30 (93.8%) | 12 (37.5%) | 25 (78.1%) |
| 7 | <5 | 4707.0 | <5 | <5 | <5 | <5 | <5 | <5 |

#### Supplementary Table 6. Treatment discontinuation and medication coverage following GLP-1RA initiation, by SMI subtype

| **Characteristic** | **Overall N = 102,041** | **No SMI N = 56,746** | **SMI N = 45,295** | **Depression N = 42,807** | **Bipolar N = 916** | **Psychosis N = 742** | **Schizophrenia N = 830** |
| --- | --- | --- | --- | --- | --- | --- | --- |
| **Treatment discontinuation, n (%)^1^** | | | | | | | |
| Ever | 56,312 (55.2%) | 31,130 (54.9%) | 25,182 (55.6%) | 23,809 (55.6%) | 491 (53.6%) | 410 (55.3%) | 472 (56.9%) |
| By 6 months | 24,928 (28.6%) | 13,826 (28.6%) | 11,102 (28.6%) | 10,452 (28.5%) | 217 (27.1%) | 206 (32.2%) | 227 (30.4%) |
| By 1 year | 34,153 (43.5%) | 18,873 (43.5%) | 15,280 (43.5%) | 14,406 (43.5%) | 300 (41.4%) | 276 (47.9%) | 298 (43.4%) |
| By 2 years | 44,064 (60.3%) | 24,340 (60.2%) | 19,724 (60.4%) | 18,624 (60.4%) | 387 (59.2%) | 335 (63.0%) | 378 (59.5%) |
| Time to first discontinuation, days, median [IQR] | 236 [63, 644] | 235 [62, 645] | 237 [67, 644] | 239 [68, 645] | 233 [68, 642] | 180 [56, 544] | 201 [56, 575] |
| Number of discontinuations, median [IQR] | 1.0 [0.0, 1.0] | 1.0 [0.0, 1.0] | 1.0 [0.0, 1.0] | 1.0 [0.0, 1.0] | 1.0 [0.0, 1.0] | 1.0 [0.0, 1.0] | 1.0 [0.0, 1.0] |
| Restarted after discontinuation, n (%)^2^ | 33,771 (60.0%) | 18,389 (59.1%) | 15,382 (61.1%) | 14,598 (61.3%) | 278 (56.6%) | 242 (59.0%) | 264 (55.9%) |
| **Proportion of days covered (PDC), %, median [IQR]^3^** | | | | | | | |
| During available follow-up | 65.9 [32.8, 94.6] | 65.0 [32.9, 94.0] | 67.0 [32.7, 95.3] | 66.8 [32.6, 95.1] | 74.8 [37.9, 96.6] | 66.2 [33.2, 96.1] | 70.3 [30.0, 96.9] |
| At 6 months | 86.3 [61.2, 98.9] | 85.2 [61.2, 98.4] | 88.0 [61.2, 99.5] | 88.0 [61.2, 98.9] | 91.8 [61.2, 100.0] | 89.1 [56.0, 99.5] | 89.3 [61.2, 100.0] |
| At 1 year | 76.7 [46.0, 97.0] | 76.7 [46.0, 96.7] | 77.3 [46.0, 97.3] | 77.0 [46.0, 97.3] | 84.4 [53.7, 98.6] | 76.7 [38.4, 97.8] | 84.7 [46.0, 98.9] |
| At 2 years | 65.9 [38.2, 94.2] | 65.2 [38.4, 93.8] | 68.5 [36.0, 94.8] | 68.1 [35.6, 94.7] | 79.0 [42.2, 96.0] | 67.0 [31.9, 95.5] | 73.7 [34.9, 97.7] |
| **High prescribing continuity (PDC >= 80%), n (%)** | | | | | | | |
| During available follow-up | 39,922 (39.1%) | 21,675 (38.2%) | 18,247 (40.3%) | 17,155 (40.1%) | 427 (46.6%) | 293 (39.5%) | 372 (44.8%) |
| At 6 months | 47,079 (55.0%) | 25,546 (53.8%) | 21,533 (56.4%) | 20,280 (56.3%) | 477 (60.5%) | 348 (55.8%) | 428 (58.5%) |
| At 1 year | 34,647 (47.4%) | 18,733 (46.4%) | 15,914 (48.7%) | 14,941 (48.5%) | 373 (55.5%) | 257 (48.0%) | 343 (53.3%) |
| At 2 years | 24,636 (40.0%) | 13,258 (39.2%) | 11,378 (41.1%) | 10,672 (40.8%) | 270 (49.2%) | 184 (41.9%) | 252 (46.3%) |
| ^1^ Discontinuation was defined as >60 consecutive uncovered days. Discontinuation by 6 months refers to treatment coverage ending before the end of 6 months post-GLP-1RA initiation, among those with follow-up at least 6 months.  ^2^ Calculated among those ever discontinued.  ^3^ PDC was calculated assuming a 28-day supply per prescription and accounting for stockpiling. Only those with follow-up through the specified post-initiation period contributed to the calculation. | | | | | | | |

#### Supplementary Table 7. Sensitivity analyses of treatment discontinuation and medication coverage following GLP-1RA initiation, by SMI subtype: (A) discontinuation defined as a gap of ≥90 days; (B) medication supply calculated from dosage information

| **Characteristic** | **Overall N = 102,041** | **No SMI N = 56,746** | **SMI N = 45,295** | **Depression N = 42,807** | **Bipolar N = 916** | **Psychosis N = 742** | **Schizophrenia N = 830** |
| --- | --- | --- | --- | --- | --- | --- | --- |
| **(A) Discontinuation defined as a gap of ≥90 consecutive uncovered days (28-day supply per prescription)** | | | | | | | |
| **Treatment discontinuation, n (%)^1^** | | | | | | | |
| Ever | 50,104 (49.1%) | 27,581 (48.6%) | 22,523 (49.7%) | 21,296 (49.7%) | 436 (47.6%) | 363 (48.9%) | 428 (51.6%) |
| By 6 months | 20,556 (23.8%) | 11,356 (23.7%) | 9,200 (23.9%) | 8,658 (23.8%) | 179 (22.5%) | 168 (26.6%) | 195 (26.3%) |
| By 1 year | 28,474 (37.0%) | 15,663 (36.8%) | 12,811 (37.2%) | 12,067 (37.1%) | 253 (35.6%) | 227 (40.6%) | 264 (39.1%) |
| By 2 years | 37,516 (53.0%) | 20,587 (52.6%) | 16,929 (53.5%) | 15,989 (53.5%) | 325 (51.2%) | 285 (55.8%) | 330 (53.5%) |
| Time to first discontinuation, days, median [IQR] | 280 [84, 734] | 280 [78, 741] | 280 [84, 728] | 280 [84, 729] | 278 [77, 740] | 225 [57, 633] | 230 [60, 661] |
| Number of discontinuations, median [IQR] | 0.0 [0.0, 1.0] | 0.0 [0.0, 1.0] | 0.0 [0.0, 1.0] | 0.0 [0.0, 1.0] | 0.0 [0.0, 1.0] | 0.0 [0.0, 1.0] | 1.0 [0.0, 1.0] |
| Restarted after discontinuation, n (%)^2^ | 26,570 (53.0%) | 14,273 (51.7%) | 12,297 (54.6%) | 11,685 (54.9%) | 215 (49.3%) | 191 (52.6%) | 206 (48.1%) |
| **Proportion of days covered (PDC), %, median [IQR]^3^** | | | | | | | |
| During available follow-up | 65.9 [32.8, 94.6] | 65.0 [32.9, 94.0] | 67.0 [32.7, 95.3] | 66.8 [32.6, 95.1] | 74.8 [37.9, 96.6] | 66.2 [33.2, 96.1] | 70.3 [30.0, 96.9] |
| At 6 months | 86.3 [61.2, 98.9] | 85.2 [61.2, 98.4] | 88.0 [61.2, 99.5] | 88.0 [61.2, 98.9] | 91.8 [61.2, 100.0] | 89.1 [56.0, 99.5] | 89.3 [61.2, 100.0] |
| At 1 year | 76.7 [46.0, 97.0] | 76.7 [46.0, 96.7] | 77.3 [46.0, 97.3] | 77.0 [46.0, 97.3] | 84.4 [53.7, 98.6] | 76.7 [38.4, 97.8] | 84.7 [46.0, 98.9] |
| At 2 years | 65.9 [38.2, 94.2] | 65.2 [38.4, 93.8] | 68.5 [36.0, 94.8] | 68.1 [35.6, 94.7] | 79.0 [42.2, 96.0] | 67.0 [31.9, 95.5] | 73.7 [34.9, 97.7] |
| **High prescribing continuity (PDC >= 80%), n (%)** | | | | | | | |
| During available follow-up | 39,922 (39.1%) | 21,675 (38.2%) | 18,247 (40.3%) | 17,155 (40.1%) | 427 (46.6%) | 293 (39.5%) | 372 (44.8%) |
| At 6 months | 47,079 (55.0%) | 25,546 (53.8%) | 21,533 (56.4%) | 20,280 (56.3%) | 477 (60.5%) | 348 (55.8%) | 428 (58.5%) |
| At 1 year | 34,647 (47.4%) | 18,733 (46.4%) | 15,914 (48.7%) | 14,941 (48.5%) | 373 (55.5%) | 257 (48.0%) | 343 (53.3%) |
| At 2 years | 24,636 (40.0%) | 13,258 (39.2%) | 11,378 (41.1%) | 10,672 (40.8%) | 270 (49.2%) | 184 (41.9%) | 252 (46.3%) |
| **(B) Discontinuation defined as a gap of ≥60 consecutive uncovered days, with medication supply calculated from dosage information (Supplementary Methods)** | | | | | | | |
| **Treatment discontinuation, n (%)^1^** | | | | | | | |
| Ever | 56,327 (55.2%) | 30,943 (54.5%) | 25,384 (56.0%) | 23,996 (56.1%) | 497 (54.3%) | 411 (55.4%) | 480 (57.8%) |
| By 6 months | 26,773 (30.7%) | 14,740 (30.5%) | 12,033 (31.0%) | 11,351 (31.0%) | 228 (28.4%) | 210 (32.9%) | 244 (32.6%) |
| By 1 year | 35,936 (45.8%) | 19,734 (45.5%) | 16,202 (46.1%) | 15,296 (46.2%) | 312 (43.0%) | 279 (48.5%) | 315 (45.7%) |
| By 2 years | 45,367 (62.1%) | 24,878 (61.7%) | 20,489 (62.7%) | 19,367 (62.7%) | 399 (61.0%) | 336 (63.3%) | 387 (60.6%) |
| Time to first discontinuation, days, median [IQR] | 204 [56, 583] | 204 [56, 583] | 204 [56, 584] | 205 [56, 583] | 215 [59, 634] | 170 [56, 532] | 178 [56, 566] |
| Number of discontinuations, median [IQR] | 1.0 [0.0, 1.0] | 1.0 [0.0, 1.0] | 1.0 [0.0, 1.0] | 1.0 [0.0, 1.0] | 1.0 [0.0, 1.0] | 1.0 [0.0, 1.0] | 1.0 [0.0, 1.0] |
| Restarted after discontinuation, n (%)^2^ | 34,574 (61.4%) | 18,669 (60.3%) | 15,905 (62.7%) | 15,096 (62.9%) | 289 (58.1%) | 249 (60.6%) | 271 (56.5%) |
| **Proportion of days covered (PDC), %, median [IQR]^3^** | | | | | | | |
| During available follow-up | 63.1 [22.7, 94.9] | 63.9 [23.0, 95.1] | 62.1 [22.4, 94.5] | 62.0 [22.3, 94.5] | 67.5 [26.8, 95.7] | 63.3 [22.6, 95.2] | 64.1 [22.3, 96.3] |
| At 6 months | 80.3 [30.6, 98.4] | 80.3 [30.6, 98.4] | 80.3 [30.6, 98.4] | 79.8 [30.6, 98.4] | 84.7 [35.0, 99.5] | 78.1 [30.6, 99.5] | 82.8 [30.6, 99.5] |
| At 1 year | 69.0 [19.2, 96.7] | 69.0 [20.3, 96.7] | 69.0 [18.6, 96.4] | 69.0 [18.1, 96.4] | 76.7 [26.2, 98.2] | 66.0 [18.6, 96.2] | 75.1 [23.0, 98.4] |
| At 2 years | 57.5 [14.8, 93.6] | 57.7 [14.8, 94.1] | 57.3 [14.2, 93.0] | 56.6 [14.2, 92.9] | 70.8 [19.2, 95.2] | 57.5 [14.8, 92.3] | 62.1 [15.3, 96.9] |
| **High prescribing continuity (PDC >= 80%), n (%)** | | | | | | | |
| During available follow-up | 39,529 (38.7%) | 22,216 (39.1%) | 17,313 (38.2%) | 16,310 (38.1%) | 382 (41.7%) | 289 (38.9%) | 332 (40.0%) |
| At 6 months | 42,894 (50.1%) | 23,820 (50.2%) | 19,074 (50.0%) | 17,974 (49.9%) | 413 (52.3%) | 305 (48.9%) | 382 (52.2%) |
| At 1 year | 31,762 (43.5%) | 17,631 (43.7%) | 14,131 (43.2%) | 13,284 (43.1%) | 321 (47.8%) | 220 (41.1%) | 306 (47.6%) |
| At 2 years | 22,749 (37.0%) | 12,699 (37.5%) | 10,050 (36.3%) | 9,423 (36.0%) | 238 (43.4%) | 163 (37.1%) | 226 (41.5%) |
| ^1^ Discontinuation was defined as >90 consecutive uncovered days in panel (A) and >60 consecutive uncovered days in panel (B). Discontinuation by 6 months refers to treatment coverage ending before the end of 6 months post-GLP-1RA initiation, among those with follow-up at least 6 months.  ^2^ Calculated among those ever discontinued.  ^3^ PDC was calculated assuming a 28-day supply per prescription in panel (A) and using dosage-derived supply in panel (B), accounting for stockpiling. Only those with follow-up through the specified post-initiation period contributed to the calculation. | | | | | | | |

### **Supplementary Figures**

#### Supplementary Figure 1. Directed acyclic graph (DAG) illustrating the causal pathways between SMI status (exposure) and GLP-1RA vs non-GLP-1RA antidiabetic prescription at the time of treatment intensification (outcome)

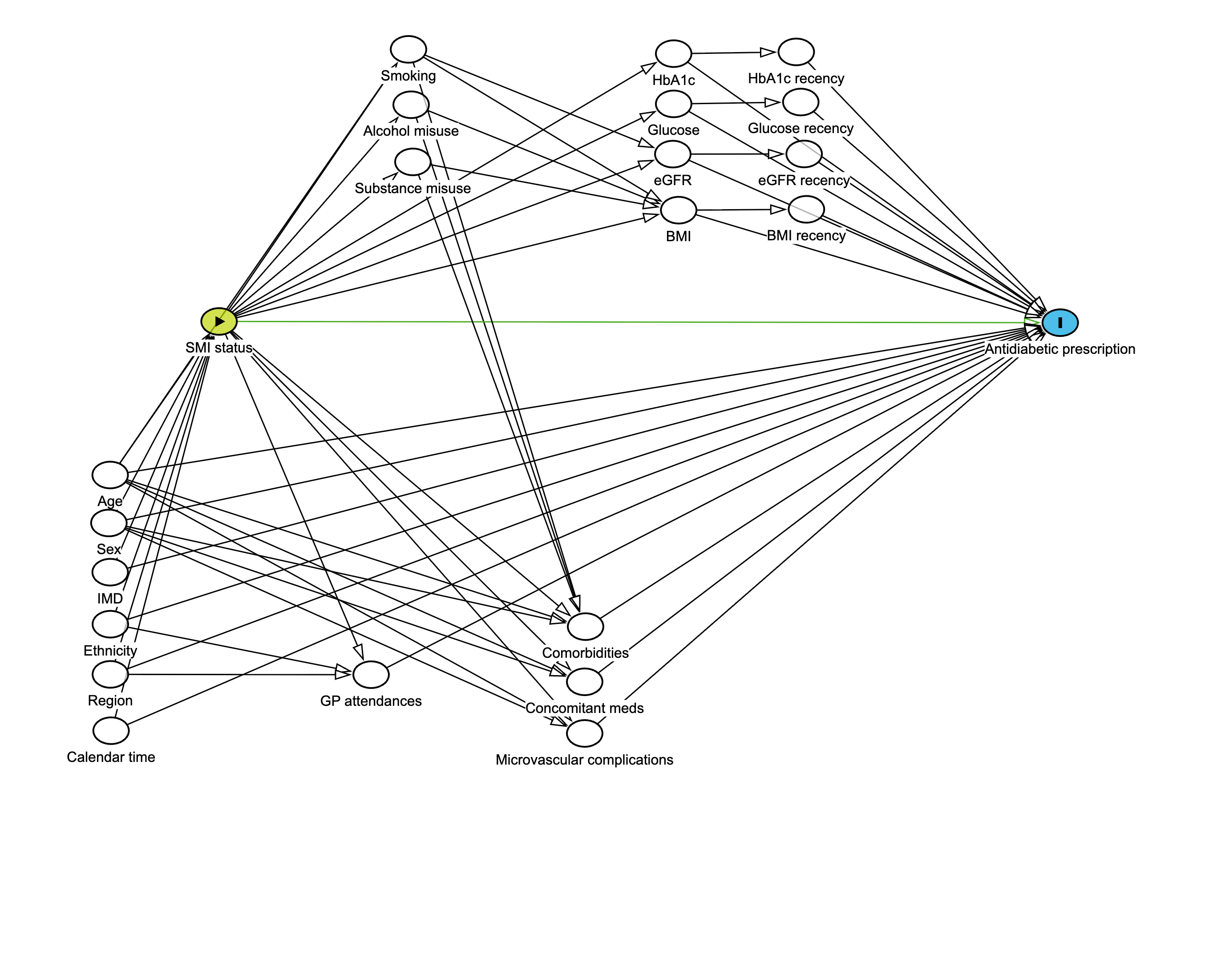

Adjustments for age, sex, social deprivation, ethnicity, region, calendar time, smoking status, alcohol misuse, substance misuse, GP attendances, microvascular complications, concomitant medications, comorbidities, biomarker (HbA1c, glucose, eGFR, BMI) and biomarker recency. The white circles represent covariates that are adjusted for. The arrows depict the direction of influence between variables. The diagram shows correct adjustment for causal identification of the direct effect of SMI status on antidiabetic prescription that does not pass through the mediating pathway of T2DM disease progression.

#### Supplementary Figure 2. Study Population CONSORT Diagram

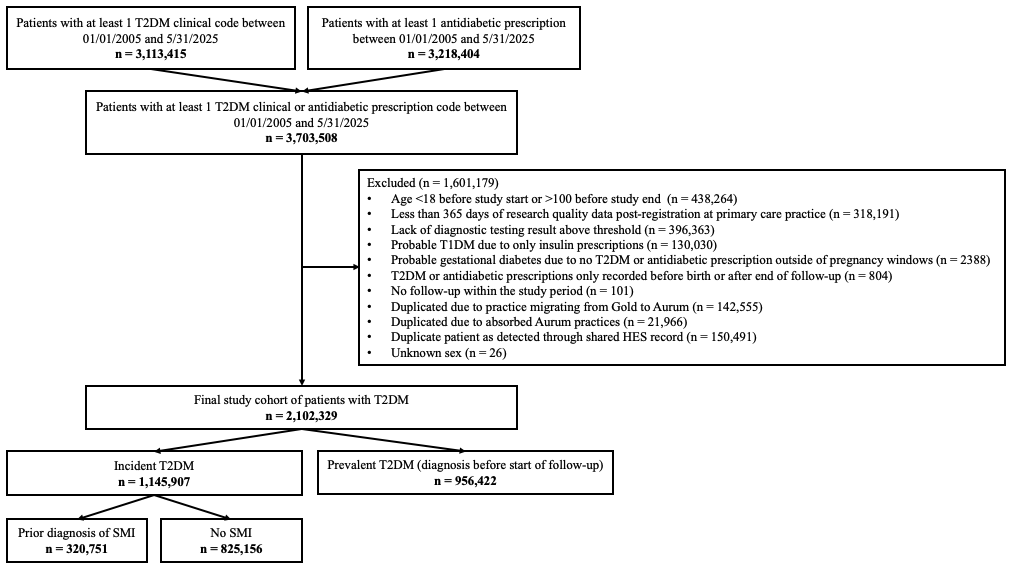

#### Supplementary Figure 3. Incidence of GLP-1RA prescribing by SMI status, among patients with incident T2DM, from 1 January 2005 to 31 May 2025.

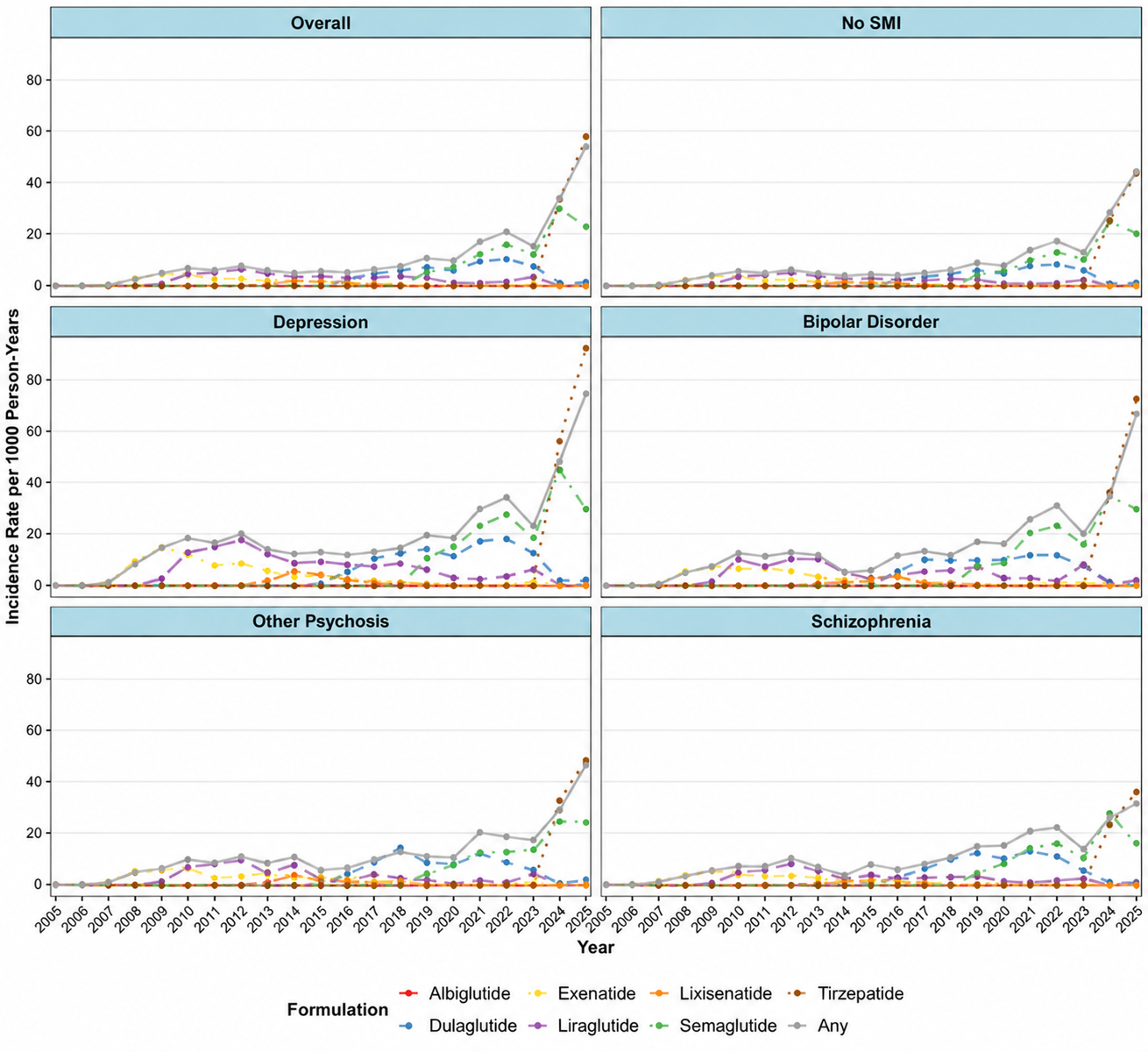

#### Supplementary Figure 4. Point prevalence rates of GLP-1RA prescribing by SMI status, among patients with incident T2DM, on 1 Feb annually from 2005 to 2025

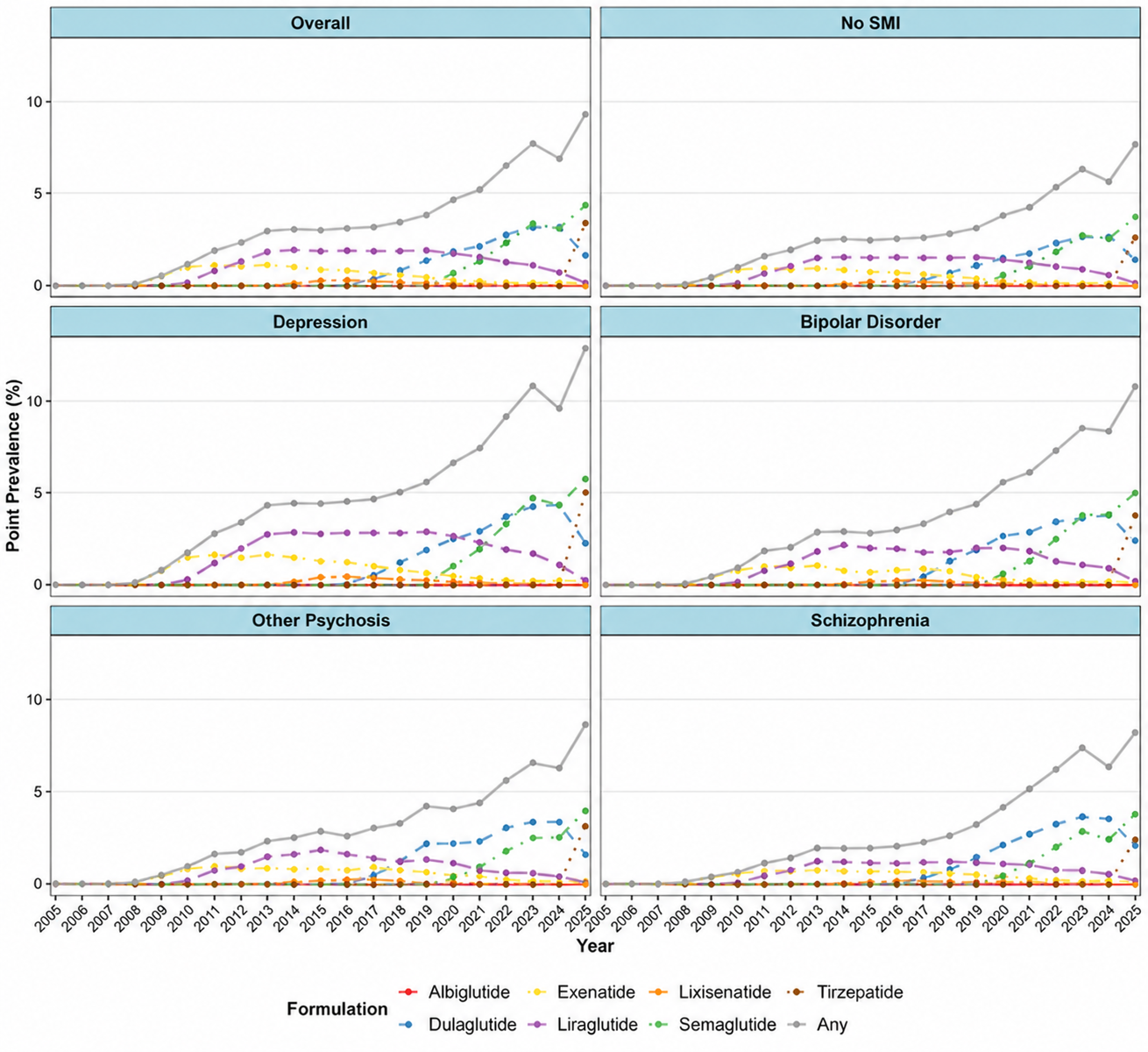

#### Supplementary Figure 5. Association between severe mental illness and initiation of GLP-1RA therapy among individuals with type 2 diabetes: complete case analysis

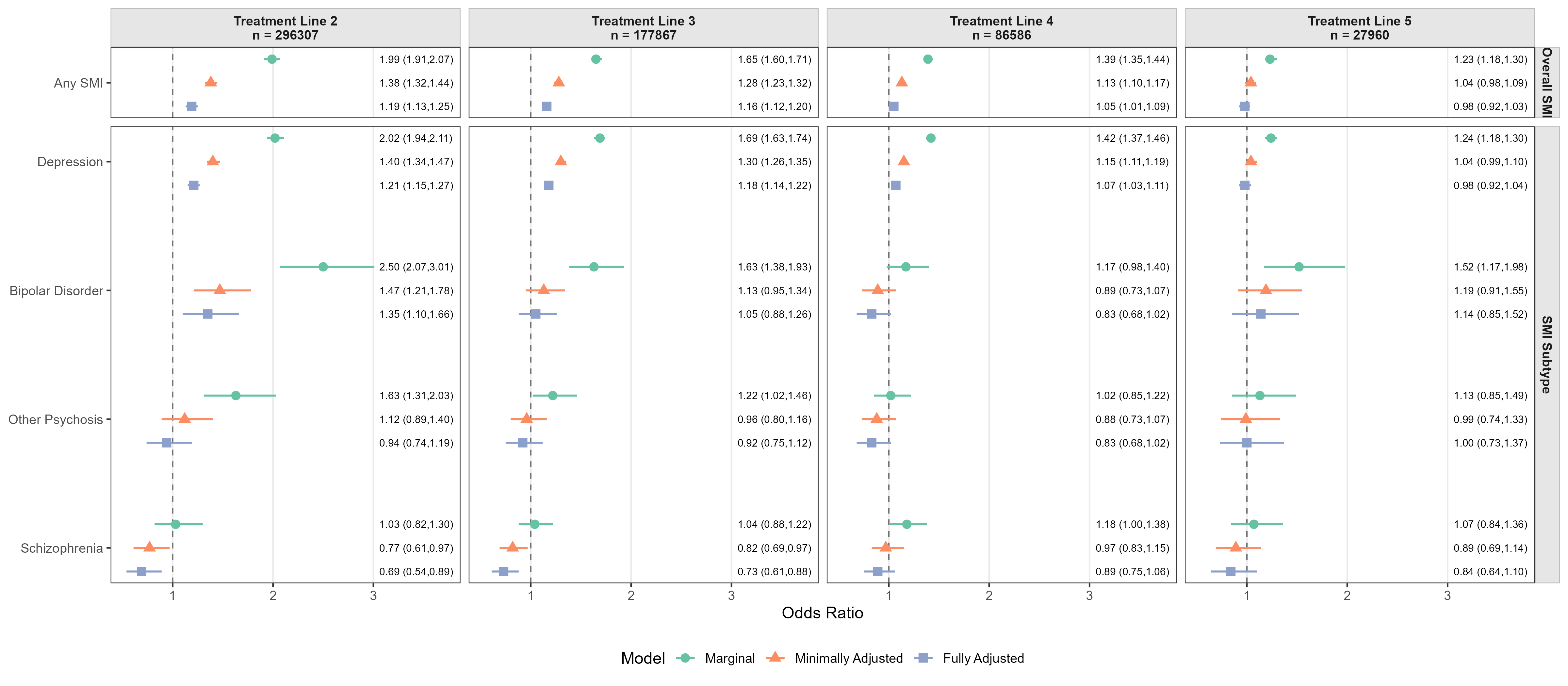

Points represent odds ratios from logistic regression models; horizontal lines represent 95% confidence intervals. The top panels show associations for any severe mental illness (SMI) versus no SMI. The bottom panels show associations for SMI subtypes, each compared with no SMI. Odds ratios greater than 1 indicate higher odds of GLP-1RA prescribing. Treatment line 2 represents second-line antidiabetic prescribing among those with prescriptions at that time; subsequent treatment lines are defined analogously. Minimally adjusted models included age, sex, region, area-level deprivation, ethnicity, and calendar year, with age centred and calendar year modelled using centred linear and quadratic terms. Fully adjusted models additionally included smoking status, alcohol misuse, substance misuse, GP attendances in the prior six months, most recent HbA1c, glucose, eGFR, and BMI, biomarker recency, hypertension, dyslipidaemia, liver disease, renal disease, cardiovascular disease, microvascular complications, and concomitant use of anticoagulants, antiplatelets, antihypertensives, and lipid-regulating medications.

#### Supplementary Figure 6. Association between severe mental illness and time to treatment intensification among individuals with type 2 diabetes

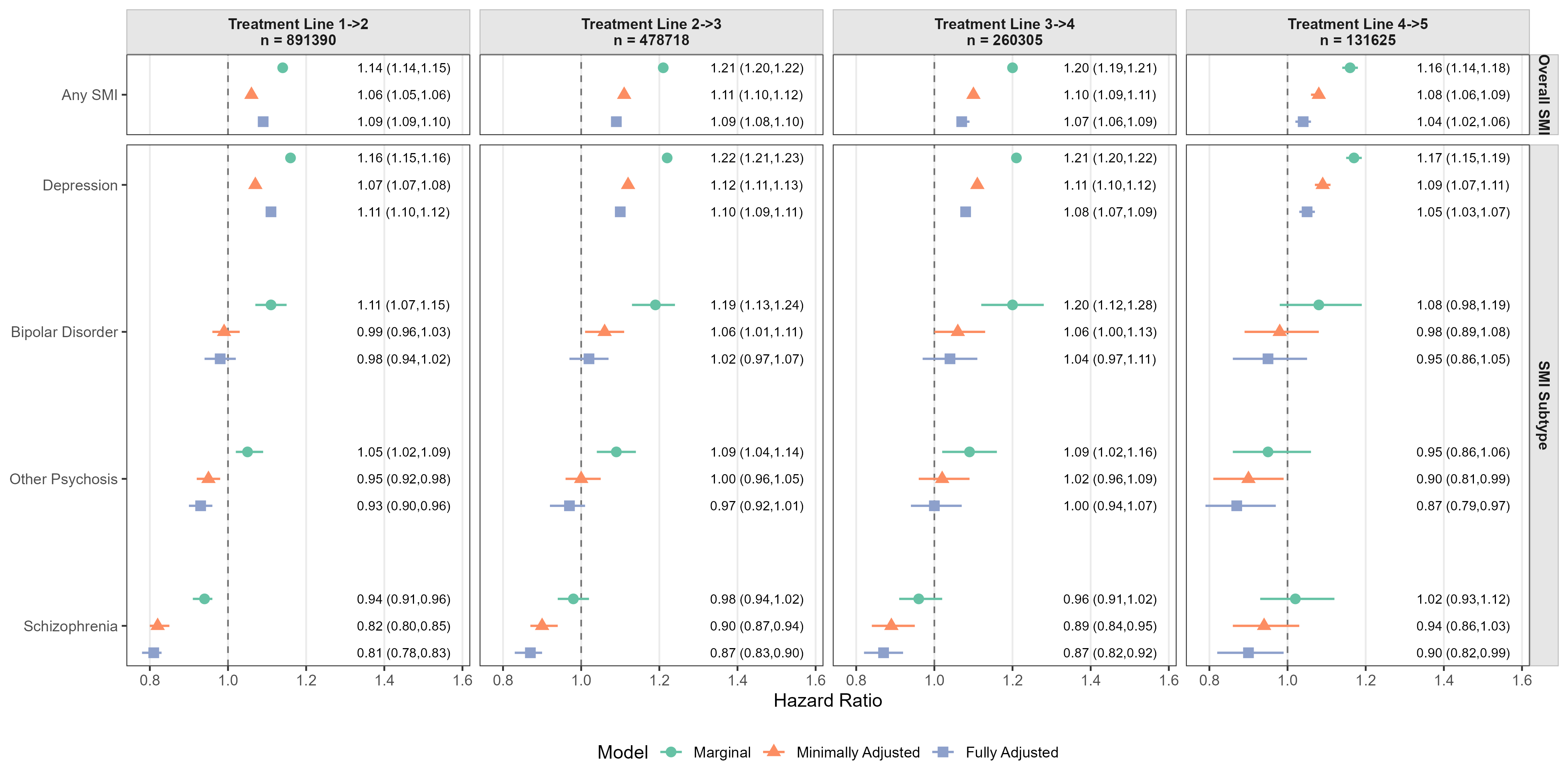

Points represent hazard ratios from Cox proportional hazards models; horizontal lines represent 95% confidence intervals. The top panels show the association for any severe mental illness (SMI) compared with no SMI. The bottom panels show associations for SMI subtypes, each compared with no SMI. Hazard ratios greater than 1 indicate faster progression to the next treatment line. Treatment line 2 represents time from initiation of first-line therapy to initiation of second-line therapy; subsequent treatment lines are defined analogously. Minimally adjusted models included age, sex, region, area-level deprivation, ethnicity, and calendar year, with age centered and calendar year modelled using centered linear and quadratic terms. Fully adjusted models additionally included smoking status, alcohol misuse, substance misuse, GP attendances in the prior six months, most recent HbA1c, glucose, eGFR, and BMI values and biomarker recency, hypertension, dyslipidaemia, liver disease, renal disease, cardiovascular disease, microvascular complications, and concomitant use of anticoagulants, antiplatelets, antihypertensives, and lipid-regulating medications.

#### Supplementary Figure 7. Association between severe mental illness and time to treatment intensification among individuals with type 2 diabetes: complete case analysis

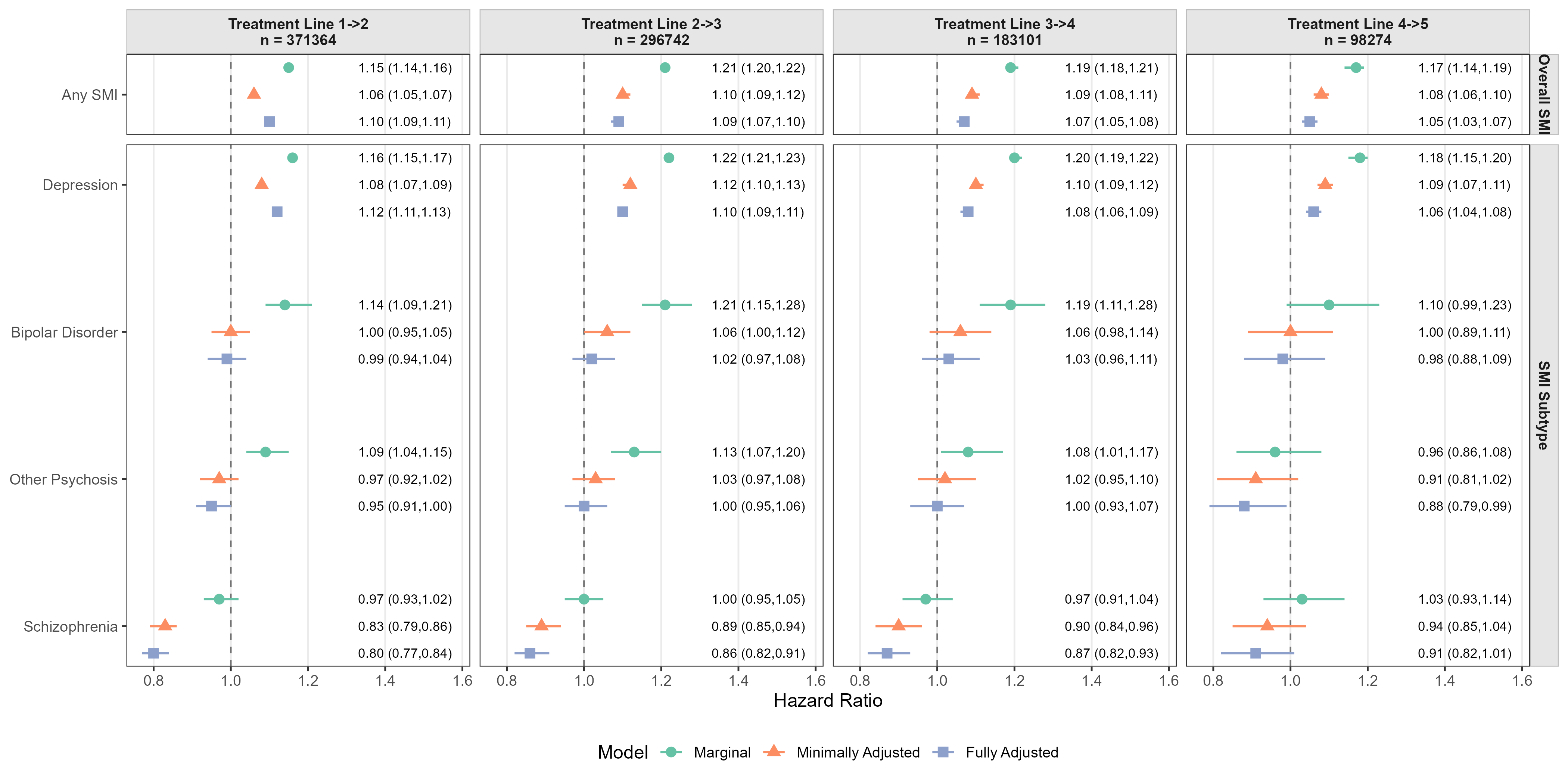

Points represent hazard ratios from Cox proportional hazards models; horizontal lines represent 95% confidence intervals. The top panels show the association for any severe mental illness (SMI) compared with no SMI. The bottom panels show associations for SMI subtypes, each compared with no SMI. Hazard ratios greater than 1 indicate faster progression to the next treatment line. Treatment line 2 represents time from initiation of first-line therapy to initiation of second-line therapy; subsequent treatment lines are defined analogously. Minimally adjusted models included age, sex, region, area-level deprivation, ethnicity, and calendar year, with age centered and calendar year modelled using centered linear and quadratic terms. Fully adjusted models additionally included smoking status, alcohol misuse, substance misuse, GP attendances in the prior six months, most recent HbA1c, glucose, eGFR, and BMI values and biomarker recency, hypertension, dyslipidaemia, liver disease, renal disease, cardiovascular disease, microvascular complications, and concomitant use of anticoagulants, antiplatelets, antihypertensives, and lipid-regulating medications.
